# Genotype-specific differences in infertile men due to loss-of-function variants in *M1AP* or *ZZS* genes

**DOI:** 10.1101/2024.03.20.24304490

**Authors:** Nadja Rotte, Jessica E.M. Dunleavy, Michelle D. Runkel, Daniela Fietz, Adrian Pilatz, Johanna Kuss, Ann-Kristin Dicke, Sofia B. Winge, Sara Di Persio, Christian Ruckert, Verena Nordhoff, Hans-Christian Schuppe, Kristian Almstrup, Sabine Kliesch, Nina Neuhaus, Birgit Stallmeyer, Moira K. O’Bryan, Frank Tüttelmann, Corinna Friedrich

**Author notes:** Correspondence: Centre of Medical Genetics, Institute of Reproductive Genetics, University of Münster, Vesaliusweg 12-14, 48149 Münster, Germany.

## Abstract

Male infertility and meiotic arrest have been linked to *M1AP*, the gene encoding meiosis I associated protein. In mice, M1AP interacts with the ZZS proteins SHOC1, TEX11, and SPO16, which promote DNA class I crossover formation during meiosis. To determine whether M1AP and ZZS proteins are involved in human male infertility by disrupting class I crossover formation, we screened for biallelic or hemizygous loss-of-function (LoF) variants in the encoding human genes to select men with a presumed protein deficiency; we compiled N=10 men for *M1AP*, N=4 for *SHOC1*, N=9 for *TEX11,* and the first homozygous LoF variant in *SPO16* in an infertile man. After in-depth characterisation of the testicular phenotype of these men, we identified gene-specific meiotic impairments: men with SHOC1, TEX11, or SPO16 deficiency shared an early meiotic arrest lacking haploid germ cells. All men with LoF variants in *M1AP* exhibited a predominant metaphase I arrest with rare haploid round spermatids, and six men even produced sporadic elongated spermatids. These differences were explained by different recombination failures: abrogated SHOC1, TEX11, or SPO16 led to incorrect synapsis of homologous chromosomes and unrepaired DNA double-strand breaks (DSB). On the contrary, abolished M1AP did not affect synapsis and DSB repair but led to a reduced number of class I crossover events. Notably, medically assisted reproduction resulted in the birth of a healthy child, offering the possibility of fatherhood to men with LoF variants in *M1AP*. Our study establishes M1AP as an important, but not essential, functional enhancer in the network of ZZS-mediated meiotic recombination.

## Introduction

Worldwide, around one in six adults is infertile (World Health Organization, 2023), and the underlying causes are equally distributed between both sexes (Vander Borght and Wyns, 2018). In men, the most severe forms of infertility are non-obstructive azoospermia (NOA) and cryptozoospermia, meaning that due to spermatogenic failure, no or only few spermatozoa are detected in the ejaculate (Nieschlag et al., 2023). For most of these cases, the only chance of fathering a child is through testicular sperm extraction (TESE) with subsequent medically assisted reproduction (MAR) using intracytoplasmic sperm injection (ICSI).

In many of these men, the absence of spermatozoa is caused by an arrest of spermatogenesis at meiosis (termed meiotic arrest or spermatocyte arrest; Wyrwoll et al., 2023b). It has been repeatedly described that this phenotype often arises via monogenic traits (Krausz et al., 2020; Wyrwoll et al., 2023a). One established disease gene for meiotic arrest is *M1AP*, which encodes meiosis 1 associated protein (Wyrwoll et al., 2020). In mice, M1AP has recently been shown to interact with three well-characterised meiosis-related proteins: SHOC1, TEX11, and SPO16 (Li et al., 2023). These form a highly conserved complex, called ZZS (from yeast Zip2/Zip4/Spo16), that in many species is crucial during prophase I of meiosis (De Muyt et al., 2018). However, it remains unknown whether the same applies for human ZZS and M1AP.

Meiosis is the crucial process during spermatogenesis that leads to the formation of haploid germ cells. A key step during meiosis is homologous recombination, which is required for accurate chromosome segregation and the formation of haploid gametes. Homologous recombination, which also enables genetic diversity, occurs via crossovers between homologous chromosomes (chiasmata). Briefly, the process is initiated by programmed DNA double-strand breaks (DSB) in early prophase I (Figure 1A). At the end of this phase, the DSBs are repaired and resolved as either non-crossovers or crossovers. When at least one crossover per homologous chromosome is formed, correct segregation can ensue; this is the *obligatory crossover* principle (repeatedly reviewed in (Bolcun-Filas and Handel, 2018; De Massy, 2013; Gray and Cohen, 2016; Xie et al., 2022)).

**Figure 1.**
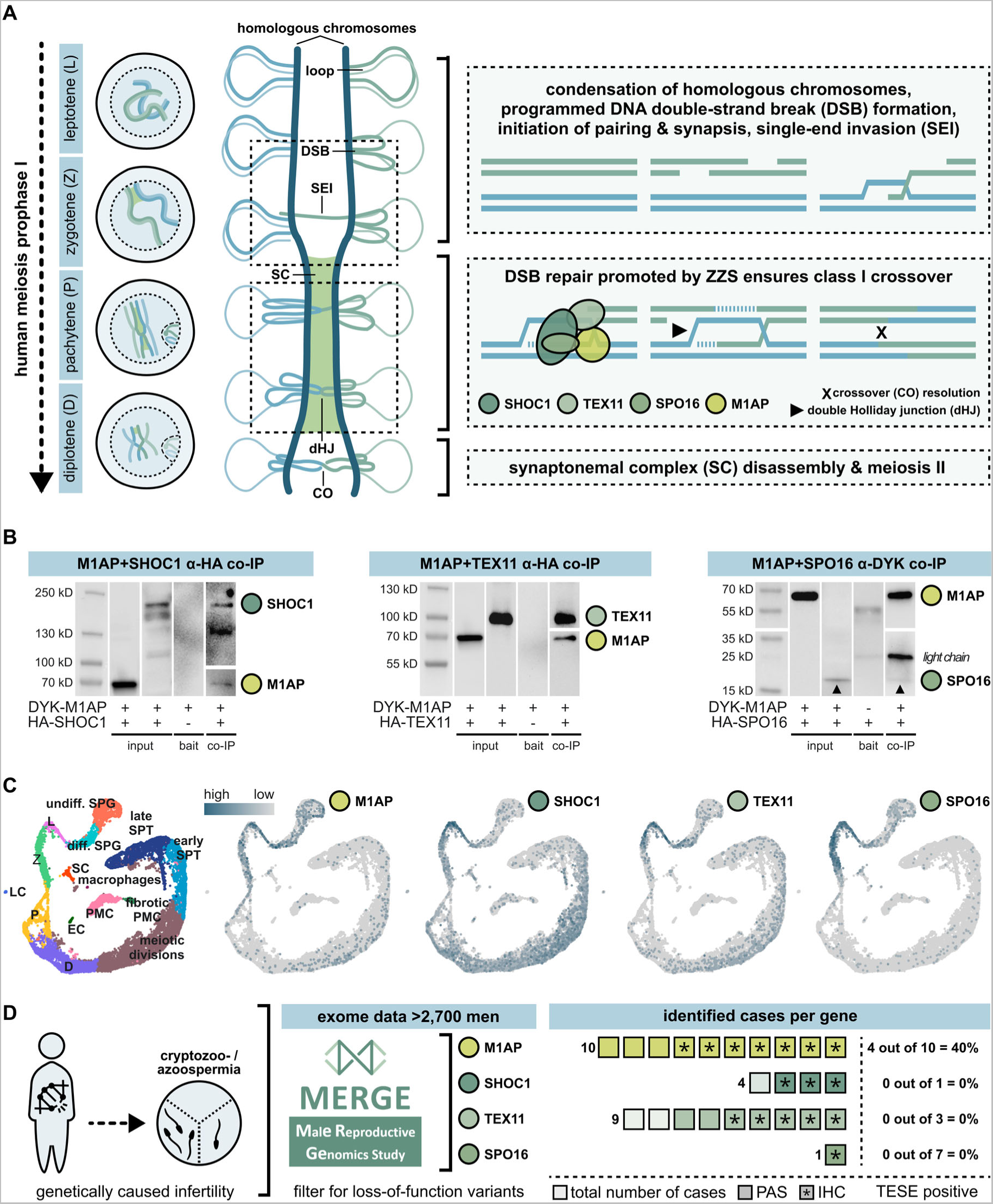
Human meiotic recombination in men depends on ZZS function – and M1AP interacts with all three complex components. A. Schematic representation of human prophase I and meiotic recombination. To simplify, the global arrangement of just one pair of homologous chromosomes is depicted and homologs are differentiated with two colours (blue, green). The molecular mechanisms of class I crossover resolution are represented in a simplified manner within the dotted boxes. In leptotene (L), homologous chromosomes duplicate, condensate, and align. DNA double-strand breaks (DSBs) are initiated. During zygotene (Z), homologous chromosomes pair up and the initiation of synapsis is supported by the dynamic assembly of a ladder-like structure – the synaptonemal complex (SC). This complex provides structural basis for meiotic recombination and in pachytene (P), the chromosomes are fully synapsed. DSB repair results in at least one crossover (CO) per chromosome pair (= obligatory crossover principle) and highly depends on SHOC1, TEX11, and SPO16 (= ZZS complex) activity. An integration of M1AP in this process was shown in mice (Li et al., 2023). Cells have completed DSB repair in diplotene (D), the SC disassembles, and homologs are physically connected by chiasmata. B. Co-immunoprecipitation (IP) proved the interaction of human M1AP (detected by N-terminal DYK-tag) with each of the ZZS complex proteins (detected by C-terminal HA-tag). Input and bait proteins served as positive and negative controls. Respective co-IP Western blot panels can be read from left to right as following: 1^st^ lane = marker, 2^nd^ = co-transfection of both plasmids, detection of M1AP in input sample, 3^rd^ = co-transfection of both plasmids, detection of ZZS in input sample, 4^th^ = transfection of bait protein only and detection after co-IP (negative control), 5^th^ = co-transfection of both plasmids, detection of both proteins from co-IP sample. In the last panel, the upturned arrow head indicates the faint bands of SPO16 for better visualisation. Due to the low detection signal of SPO16, antibody chains of the co-IP specific anti-DYKDDDDK antibody, detected with anti-mouse HRP secondary antibody, are visible too. C. Uniform manifold approximation and projection (UMAP) plot of obstructive azoospermic controls (N=3) adapted from Di Persio et al., 2021. Through mRNA expression profiling, individual stages of human spermatogenesis were clustered and visualised. Expression data of human *M1AP*, *SHOC1*, *TEX11,* and *SPO16* mRNA was compiled by querying the dataset. D. For each gene, variants leading to dysfunctional proteins were searched for in the MERGE study dataset to associate a deficient genotype with a distinct phenotype. Overall, ten men with LoF variants in *M1AP*, four men with LoF variants in *SHOC1* (N=3 from MERGE and N=1 from GEMINI), nine men with LoF variants in *TEX11*, and one man with a LoF variant in *SPO16* were selected. If possible, material aiming for testicular sperm extraction (TESE) was used for subsequent analyses including periodic acid-Schiff (PAS) or haematoxylin and eosin (H&E) staining (both highlighted in bolder colour), immunohistochemical staining (IHC = *), and spermatocyte spreads (one sample per gene of interest). A positive TESE result was only seen in men with LoF variants in *M1AP* (N=4).

In humans, there are approximately 50 crossovers per spermatocyte, which translates to one to five crossovers per pair of homologous chromosomes (Sun et al., 2005). During meiosis, meiotic recombination is mediated by a subset of highly conserved proteins of the ZMM family (an acronym for the yeast proteins Zip1/Zip2/Zip3/Zip4, Msh4/Msh5, Mer3, and Spo16; reviewed in Pyatnitskaya et al., 2019), including ZZS proteins. In particular, these proteins assemble within a meiosis-specific structure, the synaptonemal complex (SC), support the synapsis of homologous chromosomes and stabilise recombination intermediates (reviewed in Zickler and Kleckner, 2015). Only when the intermediates are stabilised by ZZM, the DSBs can be resolved as class I crossovers (Börner et al., 2004).

Even minor errors in this tightly coordinated interplay can lead to meiotic arrest and infertility (Xie et al., 2022). Accordingly, genetic variants in each of the ZZS genes have already been linked to meiotic arrest in humans, with the typical phenotypes being NOA in men and primary/premature ovarian insufficiency (POI) in women. The ZZS gene *TEX11* is a well-established X-linked gene for clinical diagnostics in male infertility (Wyrwoll et al., 2023a; Yatsenko et al., 2015); biallelic pathogenic variants in *SHOC1* lead to infertility in men and women (Krausz et al., 2020; Ke et al., 2023); and one homozygous splice region variant in *SPO16* has recently been associated with POI (Qi et al., 2023). While a strong association between *M1AP* and male infertility has been demonstrated (Wyrwoll et al., 2020; Li et al., 2023), the protein’s molecular function remains unexplored in humans.

In this study, we identified that human M1AP interacts *in vitro* with each of the ZZS proteins. Additionally, we present the first man homozygous for a loss-of-function (LoF) variant in *SPO16*. The testicular phenotype of this man and men with LoF variants in the other ZZS genes *SHOC1* and *TEX11* shared an early prophase I arrest including asynapsis and unrepaired DSBs. In contrast, LoF variants in *M1AP* led to a metaphase I arrest, where early meiotic recombination was completed while the total number of the final recombination products, the meiotic class I crossover events, was reduced. Ultimately, this leads to a predominant meiotic arrest, but fertilisation-competent spermatozoa have on rare occasions been retrieved by testicular sperm extraction. This demonstrates that M1AP is an important but not essential functional enhancer in the complex network of meiotic recombination. Collectively, these genotype-specific differences have important clinical implications, as they can be used to guide evidence-based treatment decisions or counselling of couples with male-factor infertility.

## Results

### Human M1AP interacts with the ZZS proteins SHOC1, TEX11, and SPO16 *in vitro* and shares a similar mRNA expression profile

To assess interaction between the ZZS proteins and M1AP in humans, we performed co-immunoprecipitation (IP). Human DYK-tagged M1AP was co-expressed with human HA-tagged SHOC1, TEX11, or SPO16 in HEK293T cells. Proteins were immunoprecipitated from cell lysates by tag-specific antibodies. A subsequent Western blot showed that human M1AP binds specifically to each of the three ZZS complex components (Figure 1B), showing that M1AP interacts with the ZZS complex in humans. Assuming a closely related function of all four proteins in human meiosis, we aimed to investigate whether all four genes share a similar testicular expression profile. Thus, previously published single-cell RNA sequencing data of human testicular tissue (Di Persio et al., 2021) were queried (Figure 1C). Overall, the analysis showed that all four genes are similarly expressed in all stages of meiosis with strong mRNA abundance during early prophase I (leptotene, zygotene). *SHOC1* and *TEX11* showed an additional increase of expression during later meiotic divisions.

### Composition of the study cohort with men carrying LoF variants in *M1AP* or ZZS

To compare the protein-related impact of each of the four proteins on human meiosis, we selected cases carrying LoF variants in either *M1AP* (NM_138804.4) or one of the ZZS components encoding genes, *SHOC1* (NM_173521.5), *TEX11* (NM_001003811.2), and *SPO16* (NM_001012425.2) from our Male Reproductive Genomics (MERGE) study cohort. In addition, we included one published case with a variant in *SHOC1* from the GEMINI cohort (Nagirnaja et al., 2022; G-377). Both previously published (N=10) and novel (N=14) cases were considered for functional in-depth analysis.

We compared the phenotypes of ten unrelated men with homozygous LoF variants in *M1AP*, four with biallelic LoF variants in *SHOC1*, nine with hemizygous LoF variants in *TEX11*, and one man with a homozygous LoF variant in *SPO16* (Figure 1D, Table 1). Nine of the men with a LoF variant in *M1AP* carried the recurrent pathogenic frameshift variant c.676dup, of which four had already been published and functionally analysed (Wyrwoll et al., 2020) while one man (M3609) carried a homozygous *M1AP* splice site variant (c.1073_1074+10del). This variant was predicted to undergo nonsense-mediated decay. The functional significance was analysed *in vitro* by a minigene assay which demonstrated aberrant splicing (Appendix Figure S1).

**Table 1.**
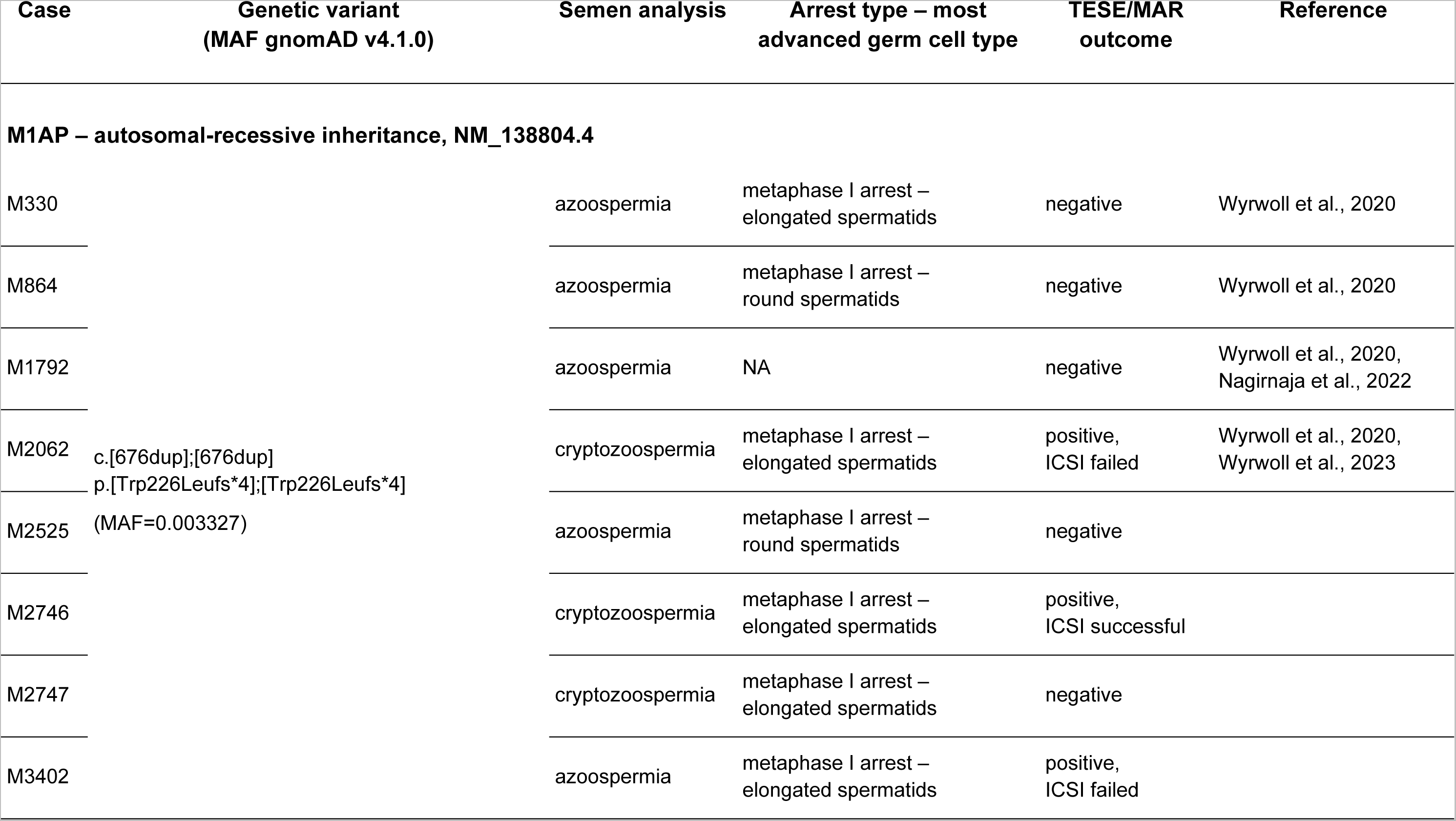
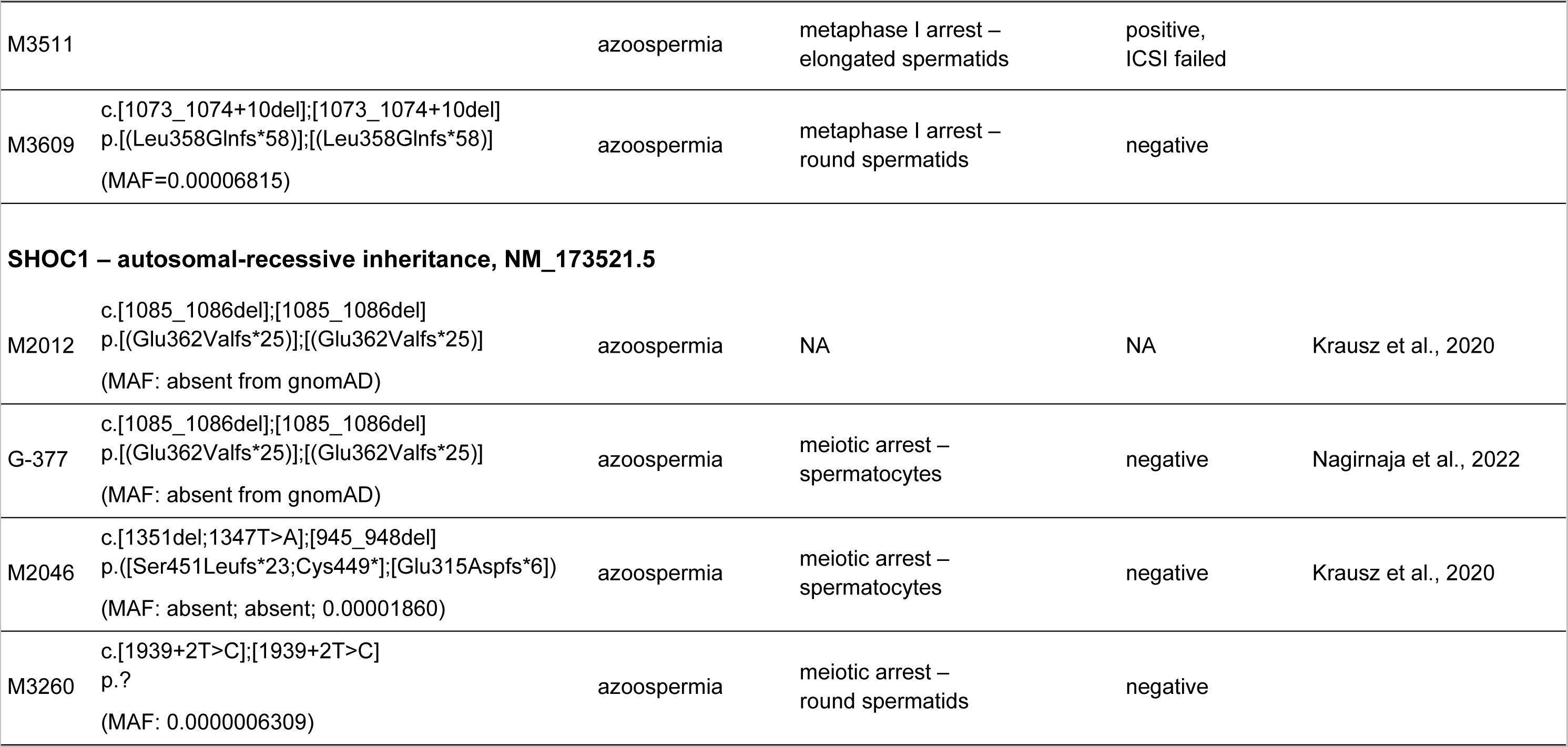
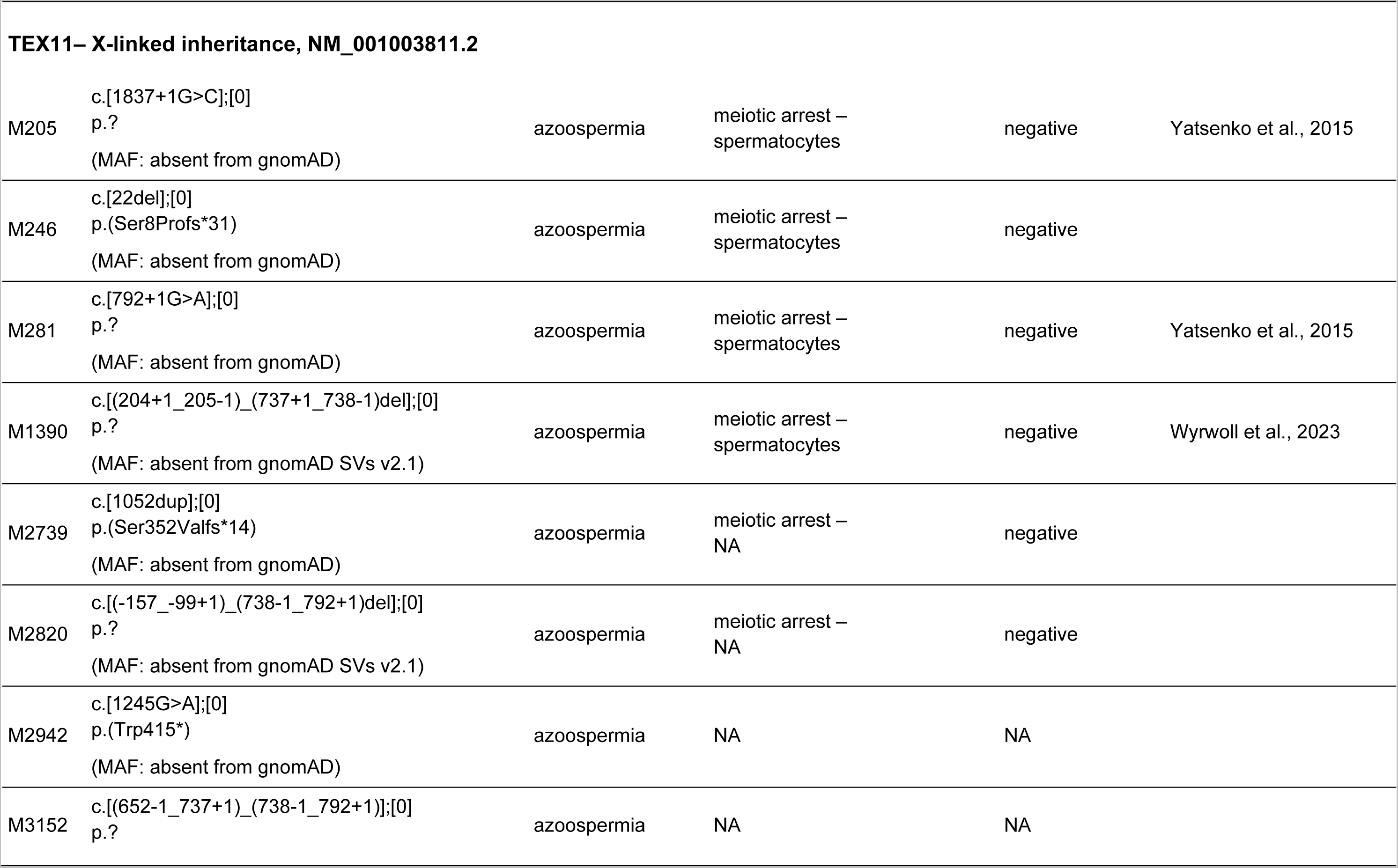
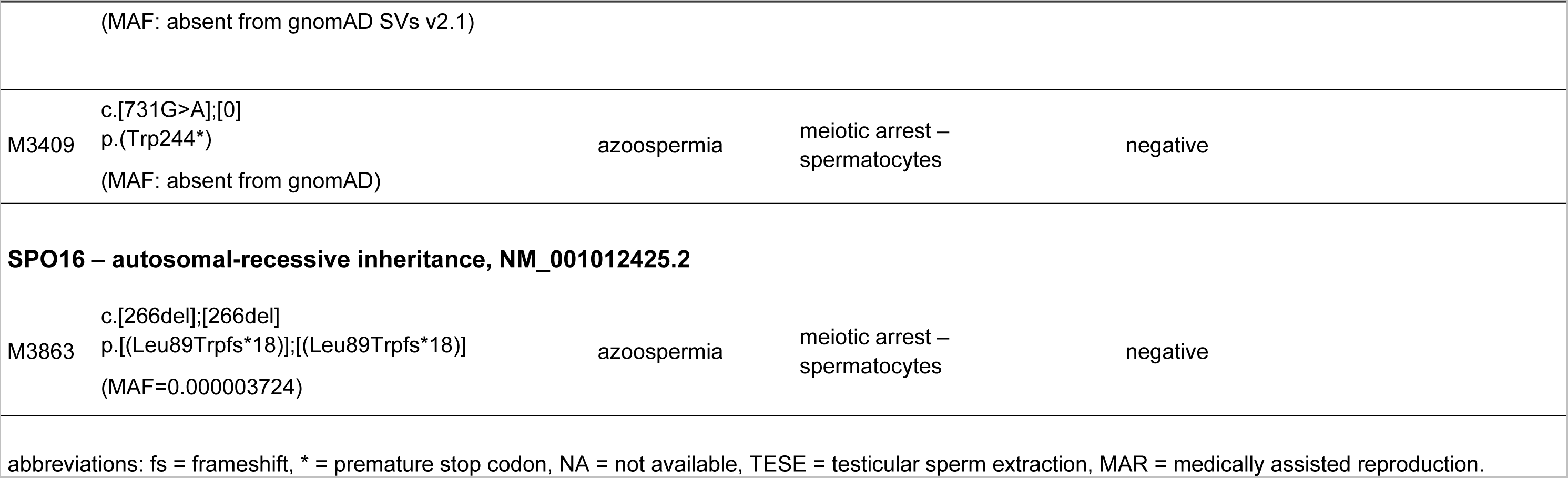
Genetic and testicular characteristics of infertile men with loss-of-function variants in *M1AP* or the ZZS genes.

For *SHOC1*, we selected two men (M2012, G-377) carrying the same homozygous frameshift variant (c.1085_1086del) and one case (M2046) with confirmed compound heterozygous variants (c.[1351del;1347T>A];[945_948del]), who were previously described (Table 1; Krausz et al., 2020; Nagirnaja et al., 2022). In addition, a novel splice site variant (c.1939+2T>C) was identified in one man (M3260). The *in vitro* minigene assay showed aberrant splicing resulting in an in-frame exon skipping event involving only ∼4% of the total protein (Appendix Figure S2) and may not lead to a complete loss of but only to a reduced protein function. This deletion affects the distant helicase hits region but not the highly conserved ‘SHOC1 homology region’, which is important for the interaction with the other ZZS proteins (Macaisne et al., 2008).

The cohort of nine men with hemizygous LoF variants in *TEX11* comprised three men with different partial gene deletions, of which two were inherited from the mother (Appendix Figures S17/S18), two men with splice site variants, and four men with variants encoding premature stop codons (Table 1); three of these cases have already been described but not in this detail (Wyrwoll et al., 2023a; Yatsenko et al., 2015).

One man (M3863) carried a novel homozygous frameshift variant (c.266del p.(Leu89Trpfs*15)) in the highly conserved ZZS gene *SPO16* (Table 1). This variant was predicted to induce a premature stop codon in exon 4 and to truncate >40% of the complete protein. Most of the protein’s central domain and the entire functional helix-hairpin-helix (HhH) domain would be affected by the truncation (Appendix Figure S3). According to the prediction, it was more likely that the mRNA was degraded by nonsense-mediated decay. In both cases, a loss of protein function can be expected.

Semen analysis of all 24 men included in this study revealed a high proportion of azoospermia (N=21), with the remaining three men displaying cryptozoospermia. Interestingly, all three cryptozoospermic men were homozygous for the recurrent LoF variant in *M1AP* (c.676dup). Clinical features were similar between men, including normal testicular volumes, normal luteinising hormone, follicle-stimulating hormone in the high range, and normal testosterone (Appendix Table S1; Appendix Figure S4).

### Men with LoF variants in *M1AP* or ZZS showed gene-specific spermatogenic defects

Testicular biopsies originating from testicular sperm retrieval (TESE) procedures were available in 21 of the selected cases and used for histological evaluation of spermatogenic differentiation. Analysis using periodic acid-Schiff (PAS, N=14) or haematoxylin and eosin (H&E, N=6) staining determined a common testicular phenotype of predominant spermatocyte arrest in all cases (Human Phenotype Ontology [HPO] term: HP:0031039, Figure 2A; Appendix Figure S5/S6). In addition, we observed cells in diakinesis or metaphase I stage characterised by scattered and misaligned chromosomes (Figure 2A, detail view). Of note, we only saw metaphase cells with correctly aligned chromosomes in controls and in men with LoF variants in *M1AP*.

**Figure 2.**
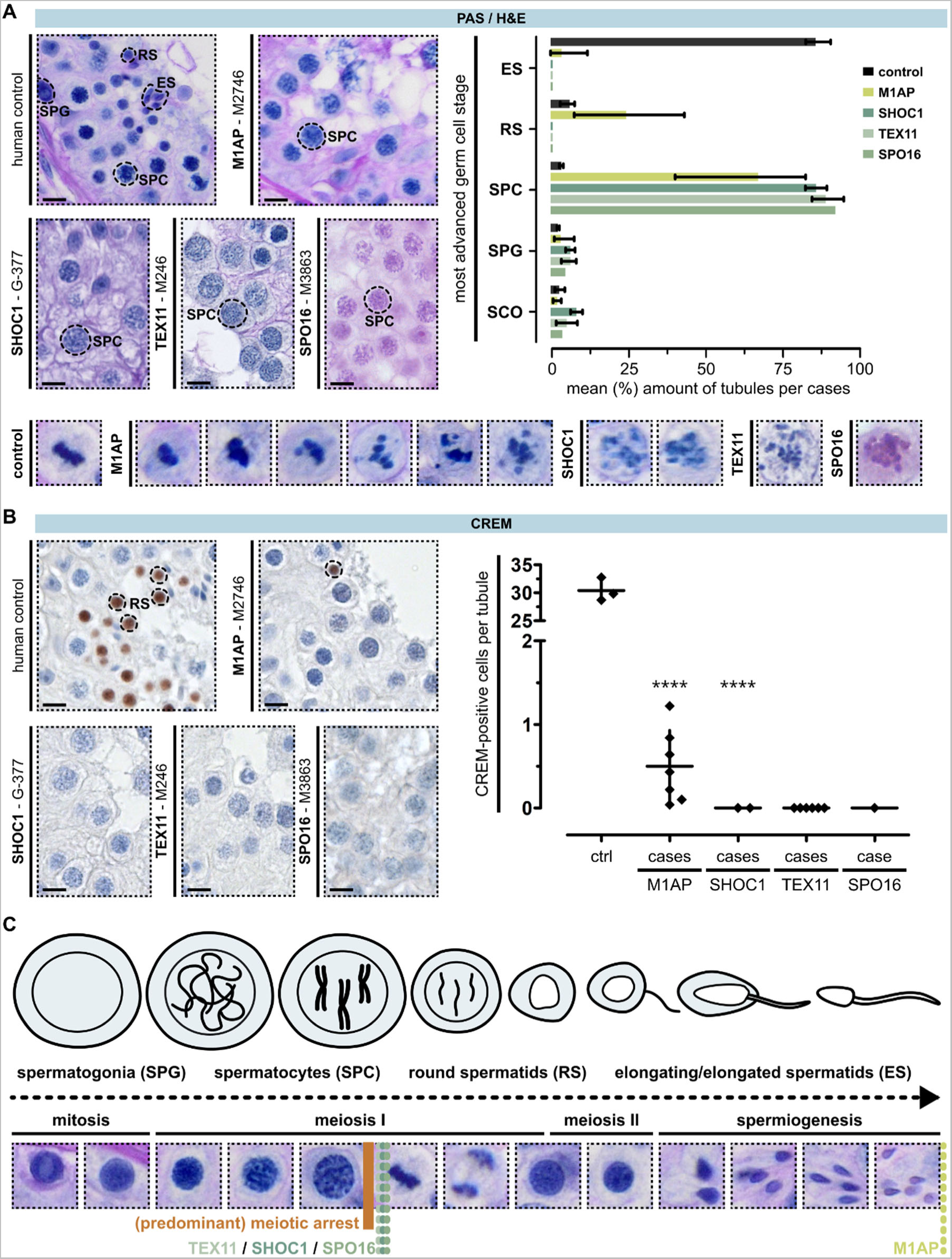
Testicular phenotyping of men with loss-of-function variants in *M1AP*, *SHOC1*, *TEX11*, and *SPO16*. A. PAS staining of testicular tissue with full spermatogenesis and infertile men with LoF variants in genes encoding the ZZS proteins and M1AP. One representative case per gene is depicted (*M1AP*: M2746, *SHOC1*: G-377, *TEX11*: M246, *SPO16*: M3863). For all biopsies, the most advanced germ cell type per tubule was assessed and is shown in the representative image and the bar graph. In addition, intact (control, M1AP) metaphase and aberrant (M1AP, SHOC1, TEX11, SPO16) metaphase-like cells were observed (detail view). ****p ≤ 0.0001 compared to controls. Haploid germ cells were analysed by CREM localisation. Only men with LoF variants in *M1AP* had CREM-positive round spermatids. Compared to the controls, the total amount of these cells was significantly reduced. C. Schematic illustration and corresponding PAS staining of human spermatogenesis. Coloured dotted lines represent the identified gene-specific germ cell arrest. SPG: spermatogonia, SPC: spermatocytes, RS: round spermatids, ES: elongated spermatids. The scale bar represents 10 µm.

To analyse the presence of post-meiotic germ cells, we performed immunohistochemical staining for cAMP responsive element modulator (CREM), a marker protein for round spermatids (Weinbauer et al., 1998). Tissue of 16 men was available for subsequent analysis (Figure 2B, Appendix Figure S7). In fertile control samples (N=3), CREM positive spermatids were observed in almost all tubules with an average 30.42 ± 1.20 spermatids per tubule. In all men analysed with LoF variants in *M1AP* (N=7), CREM-positive spermatids were observed however in significantly lower numbers than controls with an average of 0.50 ± 0.16 cell/tubule. Indeed, some but not all seminiferous tubules were found to contain round spermatids, and elongated spermatids were only observed in six of these cases. In contrast, CREM-positive round spermatids were only observed in one man with a splice site variant in *SHOC1* (M3260), which presumably not causes a loss of protein function but only an impairment of SHOC1 function enabling significantly reduced progression of spermatogenesis up to the round spermatid stage in occasional cells (Appendix Figure S2E). In total, we detected much fewer of this germ cell subtype in M3260 than in cases with LoF variants in *M1AP*. In addition, we detected no CREM-positive round spermatids in cases with LoF variants in *SHOC1* or *TEX11*. The single case with a homozygous LoF in *SPO16* (M3863) exhibited complete meiotic arrest without CREM-positive round spermatids. These results demonstrate a different testicular phenotype between men with LoF variants in *M1AP* and men affected by LoF variants in key components of the ZZS complex (Figure 2C).

### Proper chromosome synapsis and XY body formation despite M1AP deficiency

To assess whether the LoF variants in *M1AP* and in ZZS genes led to the impaired production of post-meiotic cells because of alterations in meiotic recombination, we stained patients’ testicular tissue for the two key meiosis markers γH2AX and H3S10p. Phosphorylated histone variant H2AX (γH2AX) is a marker for DSBs and was used to analyse DSB repair and correct synapsis of homologous chromosomes. In autosomes, DSBs are repaired once the homologues have successfully aligned and synapsed, and, consequently, γH2AX staining disappears. In parallel, when meiotic sex chromosome inactivation (MSCI) takes place during pachytene, a condensed chromatin domain is formed, termed the XY body. Here, γH2AX accumulates independent of meiotic recombination-associated DSBs (Fernandez-Capetillo et al., 2003). Together, the DSB repair in autosomes and the distinct formation of the XY body suggest that cells are passing through the pachytene checkpoint (Hamer et al., 2003).

Figure 3A shows the distinct γH2AX localisation during meiosis I prophase and metaphase in a human control. Men with LoF variants in *M1AP* were characterised by a qualitatively equivalent staining pattern, with quantitatively reduced amounts of pachytene- and increased amounts of zygotene-like cells (Figure 3A, detail view, bar graph, Appendix Figure S8). Interestingly, the complete repair of DSBs post-zygotene-like and the clear restriction of γH2AX to the XY body was only observed in controls and in men with LoF variants in *M1AP*. In men with LoF variants in *SHOC1*, *TEX11*, or *SPO16*, most of the cells maintained a zygotene-to-pachytene-like stage with enlarged nuclei, accumulated DSBs, and aberrant γH2AX localisation (Figure 3A, detail view, bar graph, Appendix Figure S9). Thus, an arrest in these men occurs between zygotene- to early pachytene-like (Z*/P*), referring to a human type I meiotic (Jan et al., 2018) or pachytene arrest (Enguita-Marruedo et al., 2019). Remarkably, even though cells do not proceed properly beyond zygotene-pachytene-like stage in the majority of men with LoF variants in *SHOC1*, *TEX11*, or *SPO16*, γH2AX-positive metaphase-like or diakinesis cells (M-I*) were seen in most of the cases (N=8). These cells were characterised by their spatial distribution within the tubules and scattered chromosomes, incorrectly aligned at the metaphase plate or maintained in a diakinesis stage. While similar cells were seen in men with LoF variants in *M1AP*, we also observed in parallel γH2AX-negative, intact metaphase cells with properly aligned chromosomes (N=7).

**Figure 3.**
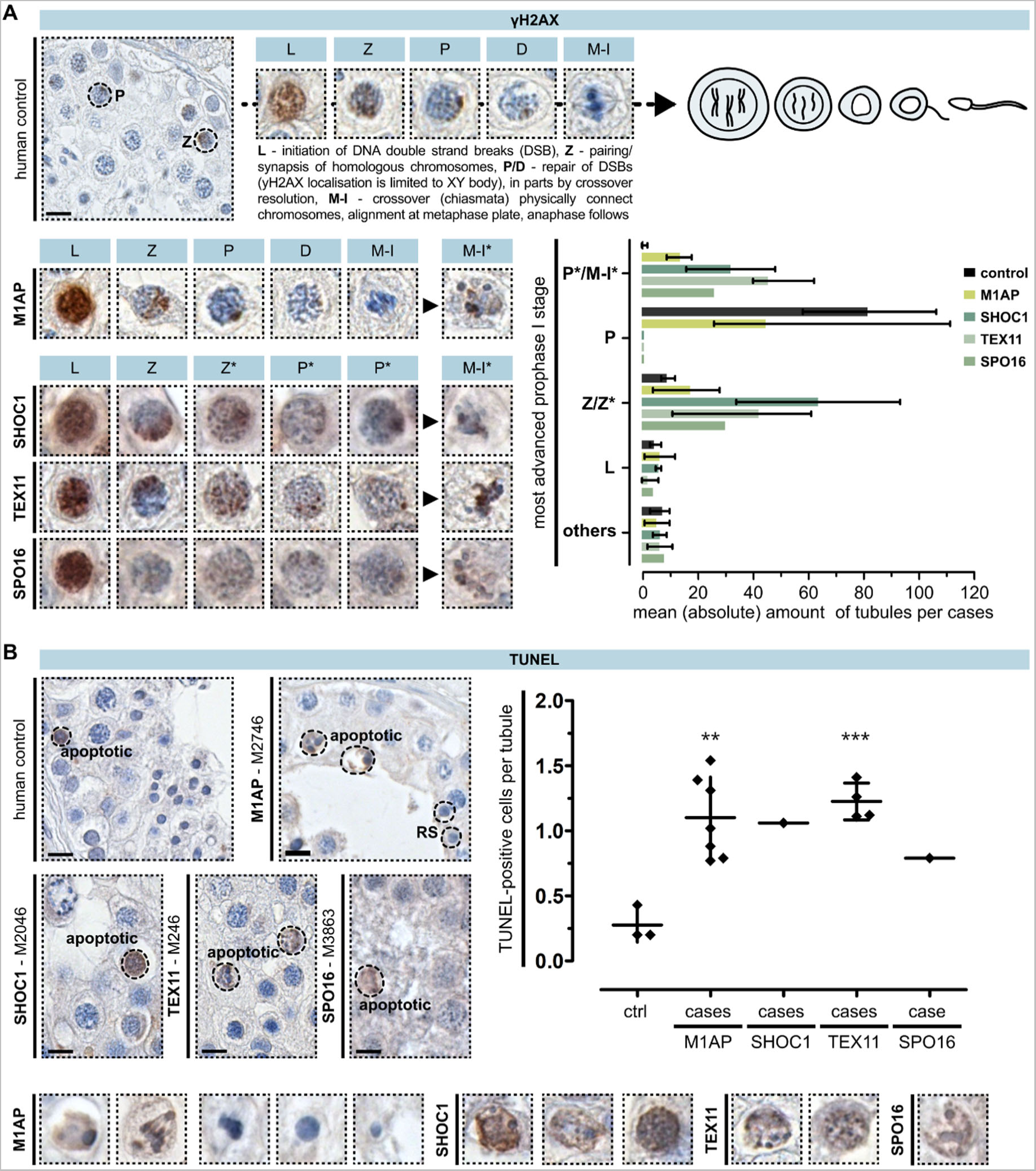
Investigation of meiotic recombination and arrest in men with loss-of-function variants in *M1AP*, *SHOC1*, *TEX11*, and *SPO16*. A. Localisation of yH2AX in a control demonstrates the marker’s specific staining pattern during each substage of meiosis prophase I. Analysis of this marker in the infertile men revealed genotype-specific aberrations and meiotic arrest: M1AP = partial metaphase (M-I) arrest, SPO16/TEX11/SHOC1 = zygotene-(Z*) to early pachytene-like (P*) arrest, with occasional metaphase-like cells (M-I*). B. TUNEL assay showed increased apoptosis in patients independent of the genetic background (dot plot). Most TUNEL-positive cells already showed hallmarks of apoptosis (detail view), and only men with LoF variants in *M1AP* showed TUNEL-negative metaphase cells with correctly aligned chromosomes, round, and elongated spermatids. **p < 0.01, ***p < 0.001 compared to controls. The scale bar represents 10 µm.

In addition, we used phosphorylated histone 3 (H3S10p) to mark cells reaching the metaphase stage (Song et al., 2011). H3 is phosphorylated when the DNA is condensed during mitosis and meiosis for subsequent segregation of the chromosomes. Localisation of H3S10p in all cases with LoF variants in *M1AP* showed that the cells with properly aligned chromosomes are meiotic metaphase cells (Appendix Figure S10). This, together with the presence of post-meiotic cells, classified the arrest in men with *M1AP* LoF variants as a partial metaphase I arrest (Enguita-Marruedo et al., 2019). In accordance with the γH2AX staining, localisation of H3S10p in men with LoF variants in *SHOC1*, *TEX11*, or *SPO16* revealed only aberrant metaphase-like/diakinesis-stage (M-I*) or mitotic spermatogonia (Appendix Figure S10).

### Increased apoptosis leads to a loss of germ cells

Maintenance of γH2AX localisation in metaphase I cells indicates a failure of meiotic recombination and checkpoint-induced apoptotic events (Enguita-Marruedo et al., 2019). Thus, we quantified apoptotic DNA fragmentation via TUNEL assay. Compared to controls, we observed a significant increase in apoptosis for all patients analysed (Figure 3B, Appendix Figure S11). Most TUNEL-positive cells showed hallmarks of apoptosis, namely condensation, fragmentation, and apoptotic bodies (Saraste and Pulkki, 2000). In addition, aberrant metaphase I-like cells with misaligned chromosomes or diakinesis-like stage cells (M-l*) were also TUNEL-positive (Figure 3B, detail view). In cases with LoF variants in *SHOC1*, *TEX11*, and *SPO16*, apoptotic DNA fragmentation was also found in rare pre-metaphase spermatocytes. One key difference between the cases was that only men with LoF variants in *M1AP* exhibited TUNEL-negative metaphase cells, round, and even elongated spermatids.

### Sufficient class I crossover events occurred despite *M1AP* LoF variants

To address whether meiotic recombination was successfully completed in spermatocytes from men with LoF variants in *M1AP*, allowing them to differentiate into post-meiotic haploid cells, we analysed spermatocyte spreads of one man (M864). In comparison, we determined the recombination failure in one man each with a LoF variant in *SHOC1* (M2046), *TEX11* (M3409), or *SPO16* (M3863). To indicate whether the synaptonemal complex (SC) formed properly, we stained for SYCP1 and SYCP3-containing elements of the SC that assemble along unaligned chromosome axes (Yuan et al., 2000). For crossover formation to occur, homologous chromosomes must undergo full synapsis, which is fulfilled when the transverse filament protein of the SC, SYCP1, localises between the axes (Dunce et al., 2018). To visualise designated crossover sites, we stained for MLH1, an endonuclease involved in the resolution of class I crossovers (Baker et al., 1996). To quantify class I crossover, we counted MLH1 foci in pachytene cells (n=10). In addition, to determine the progression of prophase I, we stained for γH2AX. Further, to analyse the correct recruitment of the ZZS complex, we labelled TEX11. For a better orientation, we marked the centromeres using a human anti-centromere antibody (ACA) (Figure 4A/B cyan, Appendix Figure S12/S13 cyan).

**Figure 4.**
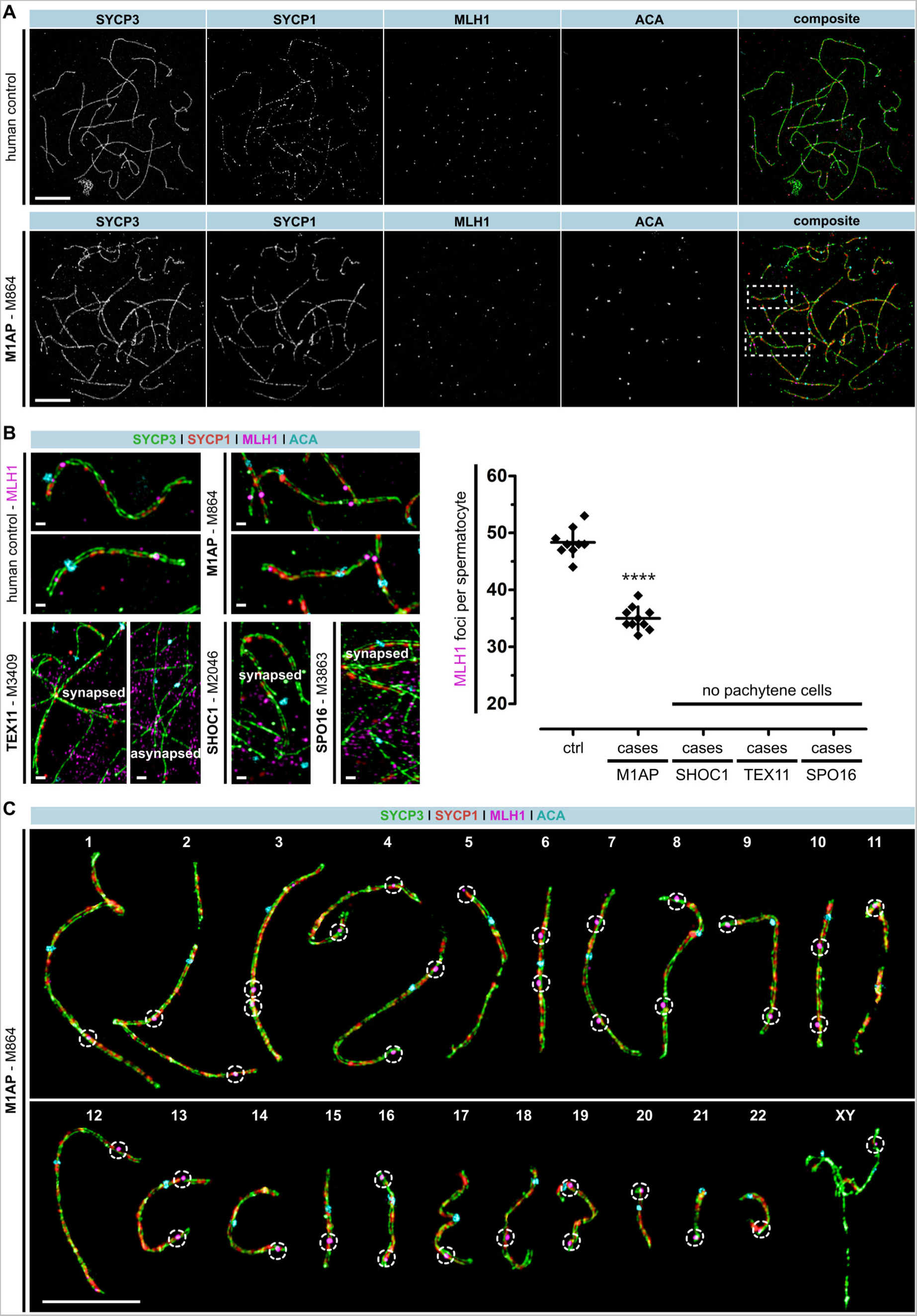
A man with loss-of-function variant in *M1AP* showed reduced MLH1 foci. A. Human spermatocyte spreads were stained for synaptonemal complex formation (= SYCP3, green + SYCP1, red), centromeric regions (= ACA, cyan), and designated crossover sites (= MLH1, magenta). A control and M864, deficient for M1AP, were compared. B. MLH1 localisation was seen on synapsed chromosome axes of pachytene-like spermatocytes in the control and M864. The total number of MLH1 foci per spermatocyte was reduced in M864. In contrast, men deficient for SHOC1 (M2046), TEX11 (M3409), or SPO16 (M8363) completely lacked pachytene-like cells. Instead, spermatocytes showed increased asynapsed chromosomes where SYCP1 localisation was lacking. No MLH1 was seen because cells arrested in a zygotene-like stage. ****p ≤ 0.0001 compared to control. C. Super resolution structured illumination microscopy (SR-SIM) of one spreaded spermatocyte. Homologous chromosomes were digitally separated to mark the MLH1 foci (dotted circles), which represent the sites of class I crossover. In this spermatocyte, each pair of homologues has at least one MLH1 focus and, thus, the *obligatory crossover* principle is met. The scale bars represent 10 µm and 1 µm for magnification.

Similar to the control, we observed spermatocytes of M864 (LoF variant in *M1AP*) in the pachytene-like stage, including properly formed chromosome axes (SYCP3) and fully synapsed homologues (SYCP1). In addition, class I crossovers were indicated by MLH1 foci (Figure 4A). In contrast, in men with SHOC1, TEX11, or SPO16 deficiency, no pachytene-like spermatocytes were identified (Figure 4B, Appendix Figure S12/S13). In all three cases, accumulated γH2AX domains were observed (Appendix Figure S13 red). In addition, SYCP1 assembly was highly reduced in M2046 (SHOC1) and facilitated to some extent in M3409 (TEX11) and M3863 (SPO16; Figure 4B red, Appendix Figure S12 red). Moreover, TEX11 was completely absent in M3409 (TEX11) and highly reduced and disorganised in M2046 (SHOC1) and M3863 (SPO16; Appendix Figure S13 magenta). In contrast, in M864 (M1AP), spermatocytes reached the pachytene-like stage, had resolved DSBs (γH2AX), and showed reduced but successful TEX11 recruitment to the chromosome axes (Appendix Figure S13 red). Importantly though, the man with a LoF variant in *M1AP* showed fewer designated crossover sites per spermatocyte (via quantification of MLH1 foci) than the control (Figure 4B magenta). Specifically, the man had on average 34.5±2 MLH1 foci per spermatocyte vs. 48±2 foci per control spermatocyte.

One crucial principle of meiotic recombination is the acquired number of crossovers per pair of homologous chromosomes, called the *obligatory crossover*. This means that each pair forms at least one crossover, crucial for the subsequent equilibrated segregation during meiosis I (reviewed in Wang et al., 2015). Taking advantage of super resolution structured illumination microscopy (SR-SIM), we investigated whether this principle was still fulfilled when M1AP was presumably dysfunctional (Figure 4C). MLH1 foci (dotted circles) were marked on each pair of homologous chromosomes from one representative spermatocyte of the man with a LoF variant in *M1AP* to localise the sites of class I crossovers. In total, 36 MLH1 foci were seen in this example, and all paired chromosomes showed at least one MLH1 focus, ensuring the obligatory crossover principle in this set of designated chromosomes.

### Men with LoF variants in *M1AP* can father healthy children by MAR

Four of the ten men with LoF variants in *M1AP* tried to conceive via ICSI. Two men (M2062 and 2746) were counselled about their genetic diagnosis before trying MAR. So far, only one man, M2746, has successfully fathered a healthy child following ICSI with ejaculated sperm. In the first and second ICSI attempts, oocyte fertilisation was possible, but cells did not develop to the blastocyst stage or did not lead to a clinical pregnancy. In the third attempt, nine oocytes were fertilised, seven of which developed up to the two-pronuclei (2N) stage. Two reached the blastocyst stage and were transferred to the female partner, after which one successfully implanted and developed normally, resulting in the birth of a healthy boy. Genome sequencing data of the child’s genomic DNA compared to the father’s demonstrated the paternity and the inheritance of the *M1AP* frameshift variant in a heterozygous state (Figure 5A). In an ICSI-TESE attempt by M2062 and his female partner, three oocytes were fertilised, but none developed to the blastocyst stage and a transfer was not possible. For M3402 and his female partner, two ICSI attempts were not successful. For M3511, five spermatozoa were retrieved by TESE and used for ICSI; however, none of the five injected oocytes were fertilised.

**Figure 5.**
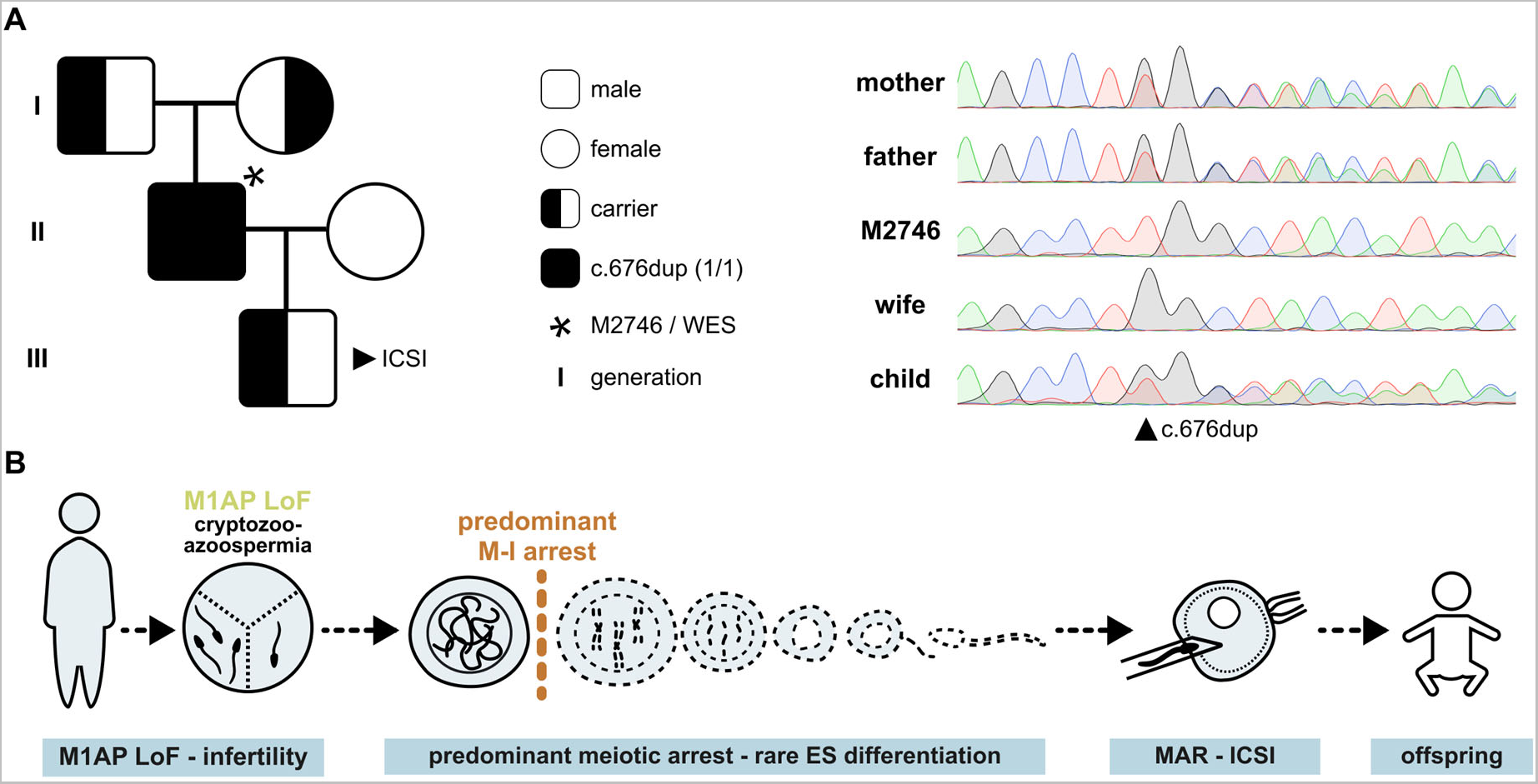
Proof-of-principle: M1AP-associated infertility can be overcome by medically assisted reproduction (MAR). A. One man (M2746) with a loss-of-function variant in *M1AP* was diagnosed with cryptozoospermia and predominant meiotic arrest. Three cycles of intracytoplasmic sperm injection (ICSI) with ejaculate-derived spermatozoa resulted in the birth of a healthy boy. Segregation analysis showed the autosomal-recessive inheritance pattern of the frameshift variant c.676dup. B. Illustration of how predominant meiotic arrest caused by M1AP-associated infertility still enables fatherhood.

To rule out any aneuploidies in the newborn boy, genome sequencing coverage data were analysed, and aberrations were excluded (Appendix Figure S14). Considering the exemplary case of M2746, ICSI with ejaculated spermatozoa or spermatozoa recovered from TESE offers the possibility of fatherhood for men with LoF variants in *M1AP* (Figure 5B).

## Discussion and conclusion

Indications that *M1AP* is required for mammalian spermatogenesis date back to 2006, when the mouse orthologue was demonstrated to be expressed in male germ cells during the late stages of spermatogenesis (Arango et al., 2006). This was strengthened by two subfertile mouse models with predominant meiotic arrest, hinting toward the protein’s role during male meiosis (Arango et al., 2013; Li et al., 2023). In addition, several human cases of male infertility have been described in which azoo-, crypto-, or extreme oligozoospermia have been associated with biallelic variants in *M1AP* (Wyrwoll et al., 2020; Tu et al., 2020; Li et al., 2023; Khan et al., 2023). Accordingly, *M1AP* has achieved a strong clinical gene-disease association based on the criteria of the Clinical Genome Resource (ClinGen) Gene Curation Working Group, and, therefore, we have proposed that this gene be included in routine diagnostics of infertile men (Wyrwoll et al., 2023a). Recently, M1AP was proposed as a fourth component of the mouse ZZS complex, linking its role to three well-known and highly conserved meiosis-related proteins: SHOC1, TEX11, and SPO16 (Li et al., 2023). In our study, we analysed the interaction between M1AP and the human ZZS proteins and showed that each ZZS protein bound to M1AP independently.

The men with LoF variants in *M1AP* investigated in this study displayed a predominant meiotic arrest with occasional haploid germ cells. Our in-depth characterisation of the patients’ testicular phenotype implied a partial metaphase I arrest, which was also seen in another reported man with a *M1AP* LoF variant (Li et al., 2023). In addition, we observed the activation of the spindle-assembly checkpoint, which is crucial for preventing premature chromosome separation and, thus, abnormal segregation and aneuploidies (reviewed in Lane and Kauppi, 2019). Accordingly, meiotic recombination was presumably intact in individual cells that progressed beyond this checkpoint. As such, an individual fertilisation-competent spermatozoon was allowed to develop, ultimately resulting in the birth of a healthy, euploid child.

These findings in humans mimic the *M1ap* knockout mouse model, which was also characterised by male subfertility (Li et al., 2023) with lower sperm counts and an increased number of arrested metaphase I cells with unaligned chromosomes. Here, fewer recombination intermediates and reduced crossover events led to metaphase I arrest and cellular apoptosis, and, consistent with the human variant carriers’ phenotype, only a fraction of cells progressed to fertility-competent spermatozoa.

In humans, *SHOC1* and *TEX11,* which encode two of the M1AP binding partners in the meiotic ZZS complex, have also previously been associated with male infertility and meiotic arrest (Krausz et al., 2020; Yatsenko et al., 2015), and both genes represent validated disease genes for male infertility (Wyrwoll et al., 2023a). In contrast to *M1AP*, hemizygous and biallelic LoF variants in these genes, respectively, have primarily been associated with a complete lack of haploid germ cells (Yatsenko et al., 2015; Krausz et al., 2020). However, the evaluation of the testicular phenotype in these initial reports was based on testicular overview staining, making a definitive conclusion difficult. In this study, we concordantly observed complete meiotic arrest and a lack of haploid, CREM-positive germ cells in all men we analysed with LoF variants in *TEX11*, pointing to an activation of the first, the pachytene, checkpoint (Yatsenko et al., 2015; Yu et al., 2021) and to an invariable genotype-phenotype correlation. This congruent phenotype was also observed in two men with LoF variants in *SHOC1.* Only in one patient with a homozygous splice site variant in *SHOC1* did we detect CREM-positive round spermatids. This splice site variant induces the in-frame deletion of a single exon. The resulting protein may still have reduced function and maintained interaction to the other ZZS proteins, due to a preserved XPF-ERCC1-like complex formation and a partly maintained distant helicase hits region, as described similarly for yeast and mouse mutants (De Muyt et al., 2018; Guiraldelli et al., 2018).

In contrast to *TEX11* and *SHOC1*, biallelic variants in *SPO16* have so far only been linked to female infertility (Qi et al., 2023); thus, our study demonstrates the first case of an infertile man with a biallelic LoF variant in *SPO16*, highlighting it as a novel candidate gene also for male infertility. His testicular phenotype and type of spermatogenic arrest is comparable to that of *TEX11* and *SHOC1* variant carriers, where no haploid germ cells were detected. Again, DSBs remain persistent and complete synapsis of chromosomes were lacking. In line, crossover resolution was absent, resulting in a complete meiotic arrest. In conclusion, in-depth testicular characterisation of the large number of men with LoF variants in *TEX11* points to a genotype-specific phenotype that correlates with the phenotype of the other two key ZZS genes, *SHOC1* and *SPO16*. However, further identification of cases is needed to fully understand how the loss of SHOC1 and SPO16 function affects meiotic progression.

Our observations on human cases are substantiated by described mouse models targeting the orthologues of the respective genes, which displayed sterility and an early spermatocyte arrest (Yang et al., 2008; Zhang et al., 2019, 2018). *Tex11* knockout mice were associated with male sterility due to apoptosis of spermatocytes, asynapsis of homologues, delayed DSB repair, and a decreased number of crossover events (Yang et al., 2008). Both complete and germ cell-specific knockout of *Shoc1* resulted in sterility. Germ cell arrest varied between zygotene and mid-pachytene stages, and cells lacked distinct XY bodies, complete chromosomal synapsis, and crossover events (Zhang et al., 2018). Homozygous knockout of *Spo16* in mice led to sterility, impaired chromosome pairing, and reduced crossover formation (Zhang et al., 2019). Compared to *Shoc1*, the *Spo16* knockout mice displayed milder defects in DSB repair and synapsis, indicating that SHOC1 alone maintains partially functionality and enables reduced DSB repair.

The common basis of all four genes *M1AP, SHOC1, TEX11*, and *SPO16*, is male infertility and meiotic arrest in the case of a loss of protein function. However, this study identified striking differences in the testicular phenotype of men with LoF variants in *M1AP* in contrast to those patients affected by LoF variants in the main ZZS complex genes *SHOC1, TEX11,* and *SPO16*. To explain why M1AP deficiency still allows a fraction of germ cells in men and mice to progress through meiosis to the haploid germ cell stage, we argue that even in the absence of M1AP, a reduced SHOC1-TEX11-SPO16 interplay is maintained: SHOC1 forms a heterodimer with SPO16 that recognises DNA-joined molecules and binds to and stabilises early recombination intermediates (Guiraldelli et al., 2018; Zhang et al., 2019). TEX11 is recruited to these sites and, in turn, assists in recruiting meiosis-specific proteins such as SYCP2 and MutSy (MSH4-MSH5), which are needed to facilitate synaptonemal complex assembly and DSB repair, finally yielding to crossover formation (Yang et al., 2008; Zhang et al., 2018).

Our findings indicate that M1AP is an important functional enhancer for promoting meiotic resolution in the ZZS network, but it is not mandatory. This is supported by the fact that although meiosis is an evolutionarily highly conserved process (reviewed in Börner et al., 2023), and, accordingly, the ZZS proteins Zip2 (SHOC1), Zip4 (TEX11), and Spo16 (SPO16) play an essential role in meiotic recombination in yeast, this lower eukaryote has no M1AP orthologue. While yeast is a well-established model organism for meiosis, M1AP is only conserved down to fish species, indicating that this protein’s function in meiotic progression is evolutionarily younger than those of the ZZS complex.

In contrast to male meiosis in mice and humans, where deficiency in M1AP or ZZS complex components results in subfertility or infertility, the role of these proteins in female fertility is less clear. In mice, female *M1ap* and *Tex11* knockouts are fertile, albeit with a reduced litter size in the case of *Tex11* knockout (Arango et al., 2013; Li et al., 2023; Yang et al., 2008). Murine *Shoc1* and *Spo16* knockouts display sterility in both sexes (Zhang et al., 2019, 2018), and in humans, biallelic LoF variants in *SHOC1* and *SPO16* have been described to lead to female infertility. For *TEX11*, only women with heterozygous variants have been found to retain their fertility (Wyrwoll et al., 2023a). For *M1AP*, however, because we and others have not identified biallelic variants in *M1AP* in fertile or infertile women, we cannot yet define the role of M1AP in human female meiosis.

*M1AP* is a clinically relevant gene for male fertility. The TESE success rate in our cohort was 40%, and we described one man who fathered a healthy, euploid child, thus showing that biallelic LoF variants in *M1AP* are compatible with fatherhood and, thereby, presenting a proof-of-principle. However, further studies are needed to fully elucidate the underlying molecular mechanisms and enable a risk estimation of checkpoint failure and aneuploidy in these men. Accordingly, it currently remains elusive whether all altered cells in men with LoF variants in *M1AP* indeed undergo checkpoint-induced apoptosis or whether rare exceptions develop independently of the spindle-assembly checkpoint resulting in aneuploid spermatozoa.

The majority of aneuploid embryos are not viable, frequently leading to spontaneous abortion (Hassold and Hunt, 2001). While studies have suggested an equal fertilisation capability between aneuploid and euploid spermatozoa, they have also evidenced a correlation between high sperm aneuploidy and recurrent ICSI or MAR failure as well as lower rates of pregnancy and live birth (reviewed in Ioannou et al., 2019). ICSI attempts with spermatozoa from two men with LoF variants in *M1AP* (M2062 and M2746) succeeded in fertilising the oocytes. Most of these did, however, not develop to the blastocyst stage or did not lead to a clinical pregnancy. Aneuploid spermatozoa could be one potential explanation for this, especially concerning chromosomes 21, 22, X, and Y, as these typically have a single recombination event and are, thus, more prone to progress without the obligatory crossover (Ferguson et al., 2007; Sun et al., 2008). This hypothesis could not be substantiated by our data, because we could not perform a direct analysis of the very few spermatozoa from any of the men to assess the frequency of aneuploidies. Thus, subsequent studies are needed to answer this question. So far, couples with M1AP-related male-factor infertility should be counselled about the small but existed chance to conceive a chromosomally-balanced biological child but also about the potential risks of aneuploidies and could benefit from preimplantation genetic testing for aneuploidy (PGT-A). Whereas the benefits of PGT-A in unexplained recurrent pregnancy failure cases remain uncertain, some studies focussing on male-factor infertility have shown improved clinical MAR outcomes after PGT-A (Rodrigo et al., 2019; Xu et al., 2021) – and M1AP-associated infertility may be such a case.

Both *M1AP* and *TEX11* represent two of the most frequently identified monogenic causes for NOA in men (Yang et al., 2015; Nagirnaja et al., 2022; Wyrwoll et al., 2023b). By identifying the first man with an underlying LoF variant in *SPO16*, we expand the clinical relevance of genetic causes to all three main ZZS genes and underline their overall importance. Here, we present the first detailed description of the testicular phenotypes of affected men, which is important for inferring the function of the proteins involved. At least for *M1AP* and *TEX11*, we provide for the first time a clear genotype-phenotype correlation, as men with LoF in the same gene show a concordant phenotype. As a result, it will now be possible to distinguish likely pathogenic variants in *TEX11* and *M1AP* from likely benign variants, i.e., those less likely to have a functional effect. Specifically, pathogenic missense variants in these genes leading to abolished protein function are expected to lead to the described, highly specific testicular phenotype. To the best of our knowledge, our study is also the first report on the birth of a healthy child (reviewed in Xie et al., 2022) having a father with a pathogenic gene variant in a clinically established disease gene for NOA. This not only distinguishes *M1AP* from other NOA genes, but it also shows that human M1AP, in contrast to the other ZZS proteins, is not required for class I crossover formation. Future studies should further address the functional links and underlying molecular details between M1AP and the human ZMM machinery.

### Material/subjects and methods Study cohort

The Male Reproductive Genomics (MERGE) study comprised exome (N=2,629) and genome sequencing (N=74) datasets of overall 2,703 probands (for details see Appendix Methods). It includes men with various infertility phenotypes: 1,622 men with azoospermia (no spermatozoa in the ejaculate, HP:0000027), 487 men with cryptozoospermia (spermatozoa only identified after centrifugation, HP:0030974), 380 men with varying oligozoospermia (total sperm count: >0 to <39 million spermatozoa, HP:0000798), 188 men with a total sperm count above 39 million, and 26 family members. Established causes for male infertility including previous radio- or chemotherapy, hypogonadotropic hypogonadism, Klinefelter syndrome, or microdeletions of the azoospermia factor (AZF) regions on the Y-chromosome were exclusion criteria. All men underwent routine physical and hormonal analysis of luteinising hormone (LH), follicle-stimulating hormone (FSH), testosterone (T) as well as semen analysis according to the respective WHO guidelines.

For this study, we selected men with rare biallelic (minor allele frequency [MAF] in gnomAD database v2.1.1, ≤0.01) or hemizygous (MAF ≤0.001) loss-of-function (LoF) variants (stop-, frameshift-, splice site variants) and deletions in *M1AP*, *SHOC1* [*C9orf84*], *TEX11*, *and C1orf146* [*SPO16*]. For consistency in the manuscript, we used the HGNC (https://www.genenames.org/) approved gene symbols for *M1AP*, *SHOC1*, and *TEX11*, whereas we refer to *C1orf146* using its alias gene symbol, *SPO16*. As a reference, the longest transcript with the highest testicular expression was chosen (*M1AP*: NM_001321739.2, *SHOC1*: NM_173521.5, *TEX11*: NM_1003811.2, *SPO16*: NM_001012425.2). If possible, segregation analysis was conducted on the DNA of family members. Samples with qualitatively and quantitatively normal spermatogenesis were included as controls (M2132, M2211, M3254, Appendix Figure S15, statistical analyses). In addition, one previously published case from an external cohort (GEMINI-377, referred to as G-377) was included for subsequent analysis (Nagirnaja et al., 2022).

### Ethical approval

All individuals gave written informed consent compliant with local requirements. The MERGE study was approved by the Ethics Committee of the Ärztekammer Westfalen-Lippe and the Medical Faculty Münster (Münster #2010-578-f-S, #2012-555-f-S; Giessen #26/11); the study of GEMINI-377 was approved by the ethics committee for the Capital Region in Denmark (#H-2-2014-103), all in accordance with the Helsinki Declaration of 1975.

### Cell culture and transient (co-)transfection experiments

Human embryonic kidney (HEK) 293T cells were cultured in Dulbecco’s modified Eagle medium (DMEM) with 10% foetal calf serum (FCS) and 1% penicillin/streptomycin (PS). The culture was maintained in T75 cell culture flasks at 37°C, 5% CO_2_, and 85% humidity.

For overexpression experiments, 400,000 cells per well were seeded in 6-well plates, cultivated for two days in DMEM-FCS without PS, and transfected at 80-100% confluence using the K2® transfection system (Biontex, #T060) according to the manufacturer’s instructions. Briefly, K2® maximiser reagent was added two hours before transfection. Directly before transfection, the medium was supplemented with 100 µm chloroquine diphosphate salt (Sigma-Aldrich, #C6628). Subsequently, 4 µg cDNA, 260 µl Opti-MEM^TM^ reduced serum medium (Gibco, #31985062), and 9 µl K2® transfection reagent per well were mixed, incubated for 15 minutes, and added to the cells. For co-transfection of DYK-tagged *M1AP* (NM_001321739.2) and either HA-tagged *SHOC1* (NM_173521.5), *TEX11* (NM_031276.3), or *SPO16* (NM_001012425.2), the total amount of DNA was maintained, and the cDNA ratio was adapted according to the respective size of each gene. Transfection and subsequent analysis were performed in three independent replicates minimum and three wells were pooled per replicate.

Cell lysis was performed 24 hours post-transfection. Accordingly, cells were washed with 1x PBS and collected in a microcentrifuge tube on ice. After centrifugation (4°C, 5 minutes, 200 rpm), cells were re-suspended with co-IP lysis buffer (25 mM Tris-HCl pH 7.4, 150 mM NaCl, 10 mM EDTA, 1% NP-40, 5% glycerol) and supplemented with 1x EDTA-free protease inhibitor cocktail (Roche, #11836170001). Samples were incubated for 15 minutes on ice and collected by centrifugation (4°C, 15 minutes, 13000 rpm). The protein-containing supernatant was transferred to a fresh microcentrifuge tube and directly processed or stored at –20°C until further usage. When co-immunoprecipitation was conducted, a small amount of lysate was kept for the ‘input’ protein validation.

### Co-immunoprecipitation

Co-immunoprecipitation protocols were optimised and, depending on the respective protein-protein interaction, two different types of magnetic beads were used for this study: Pierce^TM^ magnetic anti-DYKDDDDK tag beads (Thermo Scientific, #A36797) for M1AP-SPO16 or anti-HA tag beads (Thermo Scientific, #88838) for M1AP-SHOC1 as well as M1AP-TEX11. Beads were equilibrated and pre-cleared according to the instructions and incubated with respective protein lysates for 30 minutes at RT under gentle rotation. Anti-DYKDDDDK beads were washed with 1x PBS three times; anti-HA beads were washed with 0.025% TBS-Tween six times, and both were washed with double distilled water once. Gentle elution was achieved by incubation with a Pierce^TM^ 3x DYKDDDDK peptide (Thermo Scientific, #A36806) at 1.5 mg/ml in 1x PBS for 5 minutes shaking or by using the Pierce^TM^ elution buffer (pH 2, Thermo Scientific, #21028) for 8 minutes. Beads were collected with a magnetic stand and supernatants contained the eluted targets. Co-IP runs with pure bait lysates served as negative controls. Samples were either stored at –20°C or processed directly.

### Western blotting

Protein samples of either co-transfected DYK-*M1AP*, HA-*SHOC1*, HA-*TEX11*, or HA-*SPO16*, ‘input’ samples of co-IP eluates, or singularly transfected bait IP controls were pre-mixed 1:4 with 4x Laemmli sample buffer (Bio-Rad, #1610747) supplemented with DTT (Merck, #10197777001) and incubated at 95°C, 10 minutes for denaturation. Protein separation was achieved using 4–15% mini-PROTEAN® TGX Stain-Free^TM^ precast gels (Bio-Rad, #4568085) for SDS polyacrylamide gel electrophoresis (SDS-PAGE). Proteins were transferred to a PVDF membrane using the Trans-Blot® Turbo^TM^ mini PVDF transfer packs (Bio-Rad, #1704156EDU) following the manufacturer’s instructions. Subsequently, membranes were blocked with 5% milk powder solution in 0.025% TBS-Tween for 30 minutes at room temperature (RT). Incubation of primary antibody diluted in blocking solution was performed overnight at 4°C. Antibody details are listed in Appendix Table S3. Between the incubation steps, washing with 0.1% TBS-Tween was included. Peroxidase-conjugated secondary antibody incubation followed. Visualisation was achieved by a chemiluminescence reaction using the Clarity^TM^ Western ECL substrate kit (Bio-Rad, #1705060S) and the ChemiDoc MP imaging system (Bio-Rad, #12003154). The molecular weights of analysed proteins were calculated using a PageRuler^TM^ plus prestained protein ladder (Thermo Scientific, #26619) and image procession was performed using the Bio-Rad image lab software and molecular weight analysis tool (Bio-Rad, #12012931). Samples shown in one figure were derived from the same experiment.

### Histological evaluation of testicular biopsies

Testis biopsies of men with variants in *M1AP* or one of the ZZS genes and of control subjects were obtained from testicular sperm extraction (TESE) approaches or histological examinations at the Department of Clinical and Surgical Andrology of the CeRA, Münster, or at the Clinic for Urology, Gießen, and were included for in-depth histological phenotyping. Tissue samples were either snap-frozen, cryo-preserved (Sperm-Freeze^TM^, Ferti-Pro, #3080), or fixed in Bouin’s solution, paraformaldehyde (PFA), or GR fixative overnight. Fixed samples were washed with 70% ethanol and embedded in paraffin for routine histological examination. Tissues were sectioned at 5 µm and stained with Periodic acid-Schiff (PAS) or haematoxylin and eosin (H&E) according to standard protocols. If no differences between biopsies were observed, further analysis focused on one biopsy per case. In addition, the most advanced germ cell type was quantified, representing the percentage of elongated spermatids (ES), round spermatids (RS), spermatocytes (SPC), spermatogonia (SPG), Sertoli cell-only (SCO), and hyalinised tubules (tubular shadows, TS) per section.

### Immunohistochemical staining of human testicular tissue sections

For immunohistochemistry (IHC), biopsy samples were sectioned at 3 µm and incubated in Neo-Clear^TM^ (Sigma-Aldrich, #109843) for de-paraffinisation. Re-hydration was performed in a descending ethanol row (99%, 98%, 80%, and 70% EtOH, respectively). Between individual incubation steps, washing was performed using Tris-buffered saline (1x TBS). Incubation and washing steps were performed at RT, if not stated otherwise. Heat-induced antigen retrieval followed at 90°C, using either sodium citrate buffer (pH 6, Thermo Scientific, #005000) or Tris-EDTA buffer (pH 9, Zytomed Systems, #ZYT-ZUC029), depending on the respective antibodies’ specifications. Blocking of endogenous peroxidase activity and unspecific antibody binding was achieved by incubation in 3% hydrogen peroxidase (H_2_O_2_, Pharmacy of the University Hospital Münster, #1002187) for 15 minutes and in 25% normal goat serum (abcam, #ab7481) diluted in TBS containing 0.5% bovine serum albumin (BSA, Merck, #A9647) for 30 minutes. Primary antibodies were diluted in 5% BSA-TBS and incubated in a humid chamber at 4°C overnight, if not stated otherwise (for antibody details see Appendix Table S3). Round spermatid arrest was scrutinised by cAMP responsive element modulator (CREM) expression. Apoptosis of germ cells was analysed by TUNEL assay, following the manufacturer’s instructions (Thermo Scientific, #C10625). The progression of meiosis I was evaluated by phospho-histone H2A.X (γH2AX) staining. Metaphase cells were detected by phosphorylation of serine 10 on histone H3 (H3S10p). Incubation with unspecific immunoglobulin G (IgG) adapted to the primary antibody host system or omission of the first antibody (OC) served as negative controls (Appendix Figure S15). A secondary, biotinylated goat anti-rabbit (abcam, #ab6012) or goat anti-mouse antibody (abcam, #ab5886) was incubated for one hour, followed by conjugation with streptavidin–horseradish peroxidase (HRP, Sigma-Aldrich, #S5512) for 45 minutes. Peroxidase activity was visualised by 3,3’-diaminobenzidine tetrahydrochloride (DAB, Sigma-Aldrich, #D5905) incubation for 1– 20 minutes according to evaluation by microscope. The reaction was stopped in double distilled water. Counterstaining was performed with Mayer’s haematoxylin (Sigmal-Aldrich, #1092491000). De-hydration followed using increasing EtOH concentrations and finally, slides were cleared with Neo-Clear^TM^ and mounted with glass coverslips using M-GLAS® liquid cover glass medium (Merck, #1.03973).

### Meiotic spermatocyte spreading

Analysis of the effects of LoF variants in *M1AP*, *SHOC1*, *TEX11*, and *SPO16* on meiosis was assessed on a single-cell level by meiotic spermatocyte spreading and subsequent immunofluorescence staining. For this, snap-frozen or cryo-preserved (Sperm-Freeze^TM^, Ferti-Pro, #3080) testis samples were used (N=1 per gene [*M1AP*: M864, *SHOC1*: M2046, *TEX11*: M3409, *SPO16*: M3863]). Processing was executed at RT if not stated otherwise. All solutions were filtrated using a 0.2 µm filter. Samples were handled in 1x Dulbecco’s phosphate-buffered saline (D-PBS, Sigma-Aldrich, #14190) supplemented with 1.1 mM CaCl_2_, 0.52 mM MgCl_2_, and sodium DL-lactate solution (D-PBS^+^). Seminiferous tubules were mechanically dissected using two forceps and collected in 5 ml D-PBS^+^. Tubular remnants were allowed to settle for 5 minutes and supernatant was collected in a fresh tube. Centrifugation was performed for 5 minutes at 1000 rpm twice. The supernatant was discarded and the cell/nuclei pellet was re-suspended in D-PBS^+^ to reach 15x10^6^ cells per ml. Of this, 10 µl were mixed with 20 µl 100 mM sucrose (Thermo Scientific, #10134050) and dribbled on slides previously cleaned and dipped in freshly prepared 1% PFA (Merck, #158127, pH 9.2 set with 10 mM sodium borate, Roth, #8643) solution containing 0.15% Triton X-100 for fixation. Drying was performed for 90 minutes in a humid chamber with a closed lid and for another 45 minutes with the lid opened. Slides were washed with 0.08% Photo-Flo (Kodak, #K1464510) solution for 10 seconds and dried completely. Either, immunofluorescence staining was performed directly or slides were stored at –80°C until further processing followed.

### Immunofluorescence staining on spermatocyte spreads

Slides were defrosted for 15 minutes at RT and washed with 1x PBS three times. All solutions were filtrated using a 0.2 µm filter. The staining was divided into two parts to avoid cross reactions and minimise foci-like background. First, non-specific binding was reduced via blocking with 5% glycine (Sigma-Aldrich, #G7126), 0.3% Triton-X (Sigma-Aldrich, #T8787), and 0.01% sodium acid (NaN_3_, Sigma-Aldrich, #S2002) in PBS containing 5% normal donkey serum (Merck, #S30). Rabbit, mouse, and goat primary antibodies were diluted in 0.3% Triton-X and 0.01% NaN_3_-PBS-Tween and successively incubated overnight at 4°C, for 30 minutes at RT, and 15 minutes at 37°C in a humid chamber. Subsequently, slides were washed five times in PBS and antibodies were coupled with fluorophore-conjugated secondary antibodies for three hours at RT. Additional blocking steps followed: first in blocking solution with 5% normal donkey serum, second with 5% normal goat serum (both for 15 minutes at RT), and third by using 50 µg/ml anti-human Fab fragments in PBS (JacksonImmuno Research, #109-007-003) for 60 minutes at RT in a humid chamber. Next, a second primary antibody incubation was performed using human anti-centromere antisera (ACA, antibodiesinc, #15-234) and coupled with an anti-human fluorophore-conjugated secondary antibody for 30 minutes at RT as well as for one hour at 37°C. Finally, slides were washed five times in PBS and mounted with ROTI® mount FluorCare medium (Roth, #HP19.1). Antibody details are provided in Appendix Table S3.

### Image acquisition, processing, and digital data generation

Immunohistochemical staining was captured with an Olympus BX61VS microscope and the corresponding scanner software VS-ASW-S6, the PreciPoint O8 scanning microscope system, or a Leica DM750 microscope and the Leica ICC50 HD camera. Immunofluorescence staining of meiotic spreads was complied with a Zeiss Elyra 7 microscope for specialised 3D structured illumination (SIM^2^) and the Zeiss Zen black software. Suitable filter sets (DAPI / GFP / TXR / Y5) were used to visualise fluorophore-based antibody staining.

Image processing was achieved with the open-source software Fiji by ImageJ (v2.3.0/1.54h). For down-streaming processing of images, pictures were cropped to desired sizes to ensure a representative understanding of the testicular architecture or to allow focussed visualisation of specific details. Meiotic progression was analysed by yH2AX localisation, and all tubules of one cross section of each case were characterised by their most advanced prophase I stage. Quantification of staining was performed using the PreciPoint ViewPoint software and in-build measurement and counting tools (v1.0.0.9628). CREM- or TUNEL-positive cells were counted per round tubule per cross section and presented as average number of positive cells per tubule. Tubules were considered round when the ratio between the two diameters was in the range of 1-1.5. The minimum number of counted tubules per section was N = 25, the maximum number was N = 177. Class I crossover events were quantified for pachytene spermatocytes spreads (only possible for control and M1AP), by counting the MLH1 foci per one spermatocyte. Proper chromosome preservation was confirmed by ACA staining. Prophase I stages were confirmed by SYCP1 or yH2AX staining, depending on the respective co-labelling. Given the limited amount of material, a minimum of ten pachytene spermatocytes were analysed per case.

### Statistical analysis

Data are presented as the mean with standard deviation (SD) for scatter plots or as mean with range for bar graphs. Two-tailed Student’s unpaired t-test was applied for all statistical analyses using the GraphPad Prism software (v10.1.2). The difference was considered significant when the P value was < 0.05.

## Data availability

Sequencing data of the MERGE study is available by contacting the Institute of Reproductive Genetics (https://reprogenetik.de). Access to this data is limited for each case and specific consent of the respective samples. All novel variants have been submitted to ClinVar (SCV004708228 - SCV004708242).

## Acknowledgements

We kindly thank all probands and their families for providing data, samples, and their contents which are the foundation of our interdisciplinary biomedical research. Thomas Zobel and the team from the <u>Multiscale Imaging Centre</u>, University of Münster is thanked for his excellent and professional support in SR-SIM microscopy (Elyra Zeiss Programmnummer INST 211/901-1 FUGB). The authors thank Celeste Brennecka for language editing. In addition, we thank the following people for their valuable and professional support: Pascal Hauser, Lena Schilling, Luisa Meier, Christina Burhöi, Alexandra Hax, Jochen Wistuba, Reinhild Sandhowe, Willy Baarends, Lieke Koordnneef, Esther Sleddens, Antoine Peters, Rita Exeler, Katja Poorthuis, Adelheid Kersebom, Elke Kößer, Sophie Koser, Claudia Krallmann, and Margot Wyrwoll.

## Funding

This study was carried out within the frame of the German Research Foundation-funded Clinical Research Unit ‘Male Germ Cells’ (DFG CRU326, project number 329621271) to CF, FT, NN and the German Academic Exchange Service (DAAD) to FT and MOB (project ID 57511796).

## Conflict of interest

The authors declared no conflict of interest.

## Author contributions

All authors revised and approved the final version of the manuscript. NR and CF conceived and designed the experiments and wrote the manuscript. NR, JD, MDR, JK, AKD, SBW performed the experiments. NR, JD, SDP, CR analysed the data. VN provided the MAR data. DF, AP, NN, HCS, KA, SK, FT performed the case recruiting as well as clinical and histological evaluation. BS, MOB, FT, CF provided critical feedback and supported this work by shaping research progress and final results.

## Notes

### Competing Interest Statement

The authors have declared no competing interest.

### Author Declarations

All individuals gave written informed consent compliant with local requirements. The MERGE study was approved by the Ethics Committee of the Aerztekammer Westfalen-Lippe and the Medical Faculty Muenster: Muenster #2010-578-f-S, #2012-555-f-S; Giessen #26/11), study of GEMINI-377 was approved by the ethics committee for the Capital Region in Denmark #H-2-2014-103, all in accordance with the Helsinki Declaration of 1975.

### Summary of Updates

Text work work and improvement of figures as requested by the reviewers.

