## Supplemental file for "Genotype-specific differences in infertile men due to loss-of-function variants in *M1AP* or *ZZS* genes"

### Table of Content

**Appendix Methods**

**Appendix Tables**

Appendix Table S1. Clinical parameters of infertile men analysed in this study

Appendix Table S2. Published cases of male infertility due to LoF variants in *M1AP*, SHOC1 or *TEX11*.

Appendix Table S3. Antibody information.

Appendix Table S4. Primer information.

**Appendix Figures**

Appendix Figure S1. *M1AP* splice site variant identified in M3609.

Appendix Figure S2. *SHOC1* splice site variant identified in M3260.

Appendix Figure S3. *SPO16* loss-of-function variant identified in M3863.

Appendix Figure S4. Overview of clinical data of men with loss-of-function variants in *M1AP*, *SHOC1*, *TEX11* or *SPO16*.

Appendix Figure S5. PAS (N=7) or H&E (N=3) staining of men with LoF variants in *M1AP*.

Appendix Figure S6. PAS (N=7) or H&E (N=3) staining of men with LoF variants in *SHOC1*, *TEX11* or *SPO16*.

Appendix Figure S7. CREM staining in men with loss-of-function variants in *M1AP*, SHOC1, *TEX11* or *SPO16*.

Appendix Figure S8. γH2AX localisation showed meiosis prophase I progression in men with loss-of-function variants in *M1AP*.

Appendix Figure S9. γH2AX localisation showed impaired meiosis prophase I progression in men with loss-of-function variants in *SHOC1*, *TEX11* or *SPO16*.

Appendix Figure S10. Metaphase I cells were determined by H3S10p localisation in men with loss-of-function variants in *M1AP*, *SHOC1*, *TEX11* or *SPO16*.

Appendix Figure S11. Quantification of apoptosis in men with loss-of-function variants in *M1AP*, *SHOC1*, *TEX11* or *SPO16* via TUNEL.

Appendix Figure S12. Human spermatocyte spreads showed the absence of crossover in ZZS cases.

Appendix Figure S13. Human spermatocyte spreads showed meiotic delay in ZZS cases.

Appendix Figure S14. Euploidy analysis of M2746, his child, and the child's mother.

Appendix Figure S15. Staining of testicular tissue from a representative human control.

Appendix Figure S16. Digital droplet PCR confirmed the deletion of exons 1-11 in M2820.

Appendix Figure S17. Digital droplet PCR confirms the deletion of exons 10-11 in M3152.

**Appendix References**

### Appendix Methods

#### Exome and genome sequencing

Genomic DNA was extracted from peripheral blood leucocytes by standard methods. For exome sequencing, samples were prepared and enrichment was performed according to the protocols of either Agilent's SureSelectQXT target enrichment for Illumina multiplexed sequencing featuring transposase-based library prep technology or Twist Bioscience's twist human core exome. For library capturing, Agilent's SureSelect human all exon kits V4, V5, and V6 or Twist Bioscience's human core exome plus RefSeq spike-in and exome 2.0 plus comprehensive spike-in were used. Sample multiplexing was achieved by tagging the libraries with appropriate index primer pairs. Quality and quantity was determined using the ThermoFisher Qubit, the Agilent TapeStation 2200, and the Tecan Infinite 200Pro microplate reader. Finally, sequencing itself was performed on the Illumina HiSeq 4000, the Illumina HiSeqX, the Illumina NextSeq 500, the Illumina NextSeq 550, or the NovaSeq 6000 system, using the HiSeq 3000/4000 SBS (300 cycles), the HiSeq X Ten Reagent (300 cycles), the NextSeq 500 V2 high-output (300 cycles), or the NovaSeq 6000 S1 and S2 reagent kits v1.5 (200 cycles), respectively. Exome sequencing and analysis of patient GEMINI-377 has been described previously (Nagirnaja et al., 2022).

Genome sequencing libraries were prepared with Illumina's DNA PCR-Free library kit. Index tagging for multiplexed sequencing was accomplished by employing suitable pairs of index primers. DNA and library quantity and quality were assessed using the ThermoFisher Qubit and the Tecan Infinite 200 Pro Microplate Reader, respectively. Sequencing was performed on the NovaSeq 6000 System, utilising NovaSeq 6000 S1, S2, and S4 Reagent kits v1.5 (300 cycles), respectively.

#### Variant calling

Adapter sequences and primers were trimmed using Cutadapt v1.15 (Martin, 2011) and reads were aligned against Genome Reference Consortium human build 37 (GRCh37.p13) with BWA Mem v0.7.17 (Li and Durbin, 2010). Recalibration of base quality and variant calling was performed with the GATK toolkit v3.8 (McKenna *et al.*, 2010) or with the with Illumina Dragen Bio-IT platform v4.2, both with haplotype caller according to the best practice recommendations. Duplicate reads or reads mapping to multiple locations in the exome were excluded. Identified variants were annotated using the Ensembl variant effect predictor (McLaren et al., 2016).

**Cohort screening for high impact variants**

The MERGE sequencing data was screened for loss-of-function (LoF) (start-loss, stop-gain and frameshift) and splice site variants (± 20 nucleotides) in *M1AP*, *SHOC1* [*C9orf84*], *TEX11*, and *C1orf146* [*SPO16*]. For consistency in the manuscript, we used the HGNC (https://www.genenames.org/) approved gene symbols for *M1AP*, *SHOC1*, and *TEX11*, whereas we refer to *C1orf146* using its alias symbol, *SPO16*. We considered only variants corresponding to the respective mode of inheritance for each gene (*M1AP*/*SHOC1*/*SPO16* = autosomal recessive (AR), *TEX11* = X-linked recessive (XR)). Resulting variants were filtered for the general population frequency (gnomAD database v2.1.1; (Karczewski et al., 2020; minor allele frequency [MAF] ≤0.01 (AR) or ≤0.001 (XR)) and an occurrence ≤30 times in our in-house database containing 4,377 datasets from individuals with other genetic diseases. As reference, the longest transcript per gene with the highest testicular expression (according to GTEx; (Yu et al., 2011) was selected (*M1AP*: NM_001321739.2, *SHOC1*: NM_173521.5, *TEX11*: NM_1003811.2, and *SPO16*: NM_001012425.2). Nonsense-mediated decay prediction was performed by (https://www.mutationtaster.org/).

The sequencing data was analysed for second hits present in a list of 18 azoospermia genes with at least moderate clinical validity (Wyrwoll et al., 2023) and 363 candidate genes with strong expression in human male germ cells and an associated Gene Ontology classification of *male infertility* in the Mouse Genome Informatics Database.

**Variant validation and segregation analyses**

Identified SNVs were confirmed by Sanger sequencing. Similarly, DNA from family members was analysed for co-segregation attempts and to show if variants occur biallelic. Briefly, the region of interest was amplified using respective primers listed in Appendix Table S3. Purification of PCR products and sequencing was performed according to standard protocols.

For validation of deletions, droplet digital PCR (ddPCR) was used as described previously (Dicke et al., 2023). In brief, ddPCR was carried out using the QX200 droplet digital PCR system (Bio-Rad, #1864001), the ddPCR supermix for probes (no dUTP) (Bio-Rad, #1863024), respective primers/probes (final concentrations: 200 nM, Appendix Table S3), and 100 ng template DNA in a final volume of 20 µl. For restriction digestion, HaeIII was used. Droplets were generated using the QX200™ droplet generator (Bio-Rad, #1864002) followed by a two-step cycling protocol, set at 95°C for 10 min, with 40 cycles of 94°C for 30 s and 58°C for 1 min, with a final extension of 98°C for 10 min and a 4°C hold. The final analysis was performed with 6-FAM- and HEX-channels using the QX200 droplet reader (Bio-Rad, #1864003). Droplet digital PCR validated the identified deletions in M2820 and M3152 (Appendix Figure S16/S17)

**Ploidy analysis from genome sequencing data of M2746's offspring**

Ploidy for autosomes and gonosomes was estimated using Illumina DragenBio-IT Platform v4.2. Read counts were normalised by dividing the median read count of each chromosome by the median read count of all autosomes. Data is depicted as $log^2$.

**Minigene splicing assay**

The functional impact of the splice site variants *M1AP* c.1073_1074+10del and *SHOC1* c.1939+2T>C was determined by *in vitro* splicing assays based on a case-specific minigene construct. Therefore, the affected region of interest was cloned with adjacent intronic sequences encompassing the variant of interest in a eukaryotic expression vector (pDESTsplice) suitable to analyse splicing events. The artificial minigene construct used in this study consists of two known exons of rat *Insulin 2*, exon 3 and 4, separated by intronic sequences. Using a two-step cloning technique results in the insertion of the region of interest into those intronic sequences separating the known exons. The region of interest was amplified from genomic DNA of the respective individual or a human male control sample with standard

PCR technique. Primers are listed in Appendix Table S4. 0.5 U/µL Phusion™ high-fidelity DNA polymerase (Thermo Scientific, # F530) was used for amplification according to the manufacturer's instructions. Cloning into a pENTR/D-TOPO® vector (Thermo Scientific, #K240020) was followed by LR recombinase reaction and gateway cloning using the Gateway™ LR clonase™ enzyme mix (Thermo Scientific, #11791020) and an ultimate pDESTsplice vector (a gift from Stefan Stamm (addgene plasmid #32484, Kishore, Khanna and Stamm, 2008). Subsequently, HEK293T cells were transiently transfected with 2 µg of wildtype or case-specific minigene DNA constructs using the K2 transfection reagent according to the manufacturer's instructions. After 24 hours of transfection, total RNA was extracted using the RNeasy plus mini kit (Qiagen, #74134) and transcribed into cDNA using the ProtoScript® II first strand cDNA synthesis kit (New England Biolabs, #E6560). Amplification was conducted with primers annealing to rat insulin exon 3 and exon 4, respectively. Finally, RT-PCR products were separated on a 2% agarose gel, visualised with the TapeStation D1000 system (Agilent, #5067-5582), cut out and extracted, and confirmed by Sanger sequencing following standard protocols.

**Protein structure predictions**

Presented protein structures (M1AP, SPO16) were predicted using AlphaFold2 from EMBL-EBI (Jumper et al., 2021; Varadi et al., 2024). Images of protein structures were adapted in colour or variant consequences and exported from the EBI server.

### Appendix Tables

#### Appendix Table S1. Clinical parameters of infertile men analysed in this study.

| Case | FSH [U/L] | LH [U/L] | Testosterone [nmol/L] | Testicular volume [mL] – right/left | Reference |
|---|---|---|---|---|---|
| **M1AP** | | | | | |
| M330 | 9 | 5.3 | 14.6 | 17/23 | Wyrwoll et al., 2020 |
| M864 | 4.7 | 1.5 | 9.6 | 19/26 | Wyrwoll et al., 2020 |
| M1792 | 7.8 | 5.1 | 10.1 | 15/15 | Nagirnaja et al., 2022; Wyrwoll et al., 2020 |
| M2062 | 3.5 | 3.6 | 18.6 | 26/23 | Wyrwoll et al., 2023, 2020 |
| M2525 | 15.4 | 7.7 | 10.2 | 22/29 | |
| M2746 | 3.5 | 2.4 | 18.5 | 18/16 | |
| M2747 | 9.7 | 3.6 | 19.9 | 8/8 | |
| M3402 | 7.2 | 5.2 | 10.7 | 15/14 | |
| M3511 | 3 | 2.1 | 8.9 | 22/13 | |
| M3609 | 3.8 | 3.7 | 17.1 | 10/10 | |
| **SHOC1** | | | | | |
| M2012 | 5.9 | 4.6 | 17.1 | 23/19 | Krausz et al., 2020 |
| G-377 | 4.9 | 7.4 | 29.6 | NA | Nagirnaja et al., 2022 |
| M2046 | 15.9 | 10.1 | 6.3 | 10/10 | Krausz et al., 2020 |
| M3260 | 4.8 | 5.2 | 11.9 | 18/13 | |
| **TEX11** | | | | | |
| M205 | 6 | 2.5 | 15.1 | 25/5* | Yatsenko et al., 2015 |
| M246 | 3.3 | 4.1 | 10.9 | 26/22 | |
| M281 | 2.9 | 2.2 | 12.8 | 20/16 | Yatsenko et al., 2015 |
| M1390 | 11.6 | 5.9 | 13.1 | 12/10 | Wyrwoll et al., 2023 |
| M2739 | 7.3 | 3.1 | 15.8 | 28/28 | |
| M2820 | 15.9 | 5.2 | 10.4 | 14/5[+] | |
| M2942 | 5.5 | 2 | 14 | 15/17 | |
| M3152 | 5.3 | 3.6 | 22 | 11/10 | |
| M3409 | 7.1 | 5.3 | 8.9 | 10/10 | |

| | | | | |
|---|---|---|---|---|
| **SPO16** | | | | |
| M3609 | 5.7 | 4.1 | 14.2 | 11/13 |

abbreviations: FSH = follicle-stimulating hormone, LH = luteinising hormone, NA = not available. All parameters were obtained at the first visit. *diagnosed and treated varicocele at the age of 18, led to testis atrophy (left side), [+]maldescended testis (left side).

reference values: FSH = 1-7 IU/L, LH = 2-10 IU/L, T = >12 nmol/L, TV = >12 mL per testis (right/left).

**Appendix Table S2. Published cases of male infertility due to LoF variants in *M1AP*, *SHOC1* or *TEX11*.**

| Case # | Case ID | LoF variant | | Genotype | Phenotype | Transcript | Reference |
|---|---|---|---|---|---|---|---|
| **M1AP** | | | | | | | |
| 1 | M330 | c.676dup | p.Trp226Leufs*4 | 1/1 | azoo (MeiA – SPC), TESE negative | NM_138804.4 | Wyrwoll et al., 2020 |
| 2 | M864 | | | | azoo (MeiA – SPC), TESE negative | NM_138804.4 | Wyrwoll et al., 2020 |
| 3 | M1792 | | | | azoo (MeiA – SPC), TESE negative | NM_138804.4 | Nagirnaja et al., 2022; Wyrwoll et al., 2020 |
| 4 | M2062 | | | | crypto (MeiA – ES) | NM_138804.4 | Wyrwoll et al., 2023, 2020 |
| 5 | RU01691 | | | | azoo (MeiA – postmeiotic cells), TESE positive | NM_138804.4 | Wyrwoll et al., 2020 |
| 6 | MI-0006-P | | | | azoo (MeiA – RS [note: not validated by CREM IHC]), TESE negative | NM_138804.4 | Wyrwoll et al., 2020 |
| 7 | F1: II-1 | c.1435-1G>A | p.? | 1/1 | severe oligozoospermia | NM_138804 | Tu et al., 2020 |
| 8 | | c.1074+2T>C | p.Ala312Lysfs*7 | 1/1 | azoo (M-I arrest – RS [note: not validated by CREM IHC]) | NM_001321739.2 | Li et al., 2023 |
| 9 | 9-azoo | c.(*142767)_(*153120 )del | p.? | 1/1 | azoo | NM_001281296.2 | Khan et al., 2023 |

| | | | | | | | |
|---|---|---|---|---|---|---|---|
| 10 | GEMINI-1678 | c.676dup | p.Trp226Leufs*4 | 1/1 | severe oligozoospermia | NM_001281296.2 | Khan et al., 2023 |
| 11 | GEMINI-283 | | | | NOA | NM_001281296.2 | Khan et al., 2023 |
| **SHOC1** | | | | | | | |
| 1 | 11-272 | c.797del | p.(Leu266Glnfs*6) | 1/1 | SPC arrest, M-I | NM_173521.4 | Krausz et al., 2020 |
| *2* | M2046 | c.[1351del;1347T>A];[945_948del] | p.([Ser451Leufs*23;Cys449*];[Glu315Aspfs*6]) | 1/1 | MA | NM_173521.4 | Krausz et al., 2020; this study |
| 3 | M2012 | c.1085_1086del | p.(Glu362Valfs*25) | 1/1 | azoo (MA) | NM_173521.4 | Krausz et al., 2020; Wyrwoll et al., 2023; this study |
| *4* | Family 1 | c.1582C>T c.231_232del | p.(Arg528*) p.(Leu78Serfs*10) | 1/1 | MA (SPC) | NM_173521 | Yao et al., 2021 |
| 5 | Family 2 | c.1194del | p.(Leu400Cysfs*8) | 1/1 | MA | NM_173521 | Yao et al., 2021 |
| *6* | sporadic MA | c.1464del | p.(Asp489Thrfs*14) | 1/1 | MA | NM_173521 | Yao et al., 2021 |
| *7* | F1:II-2 | c.231_232del | p.(Leu78Serfs*10) | 1/1 | MA (SPC) | NM_173521 | Wang et al., 2022 |
| 8 | F2:II-1 | | | | MA (SPC) | NM_173521 | Wang et al., 2022 |
| *9* | GEMINI-377 | c.1085_1086del | p.(Glu362Valfs*25) | 1/1 | NA | NM_173521.4 | Nagirnaja et al., 2022; this study |

**TEX11**

| | | | | | | | |
|---|---|---|---|---|---|---|---|
| 1 | Patient 1 | c.(651+1_652-1)_(888+1_889-1)del | | 1/- | mixed testicular atrophy | NM_001003811 | Yatsenko et al., 2015 |
| 2 | Patient 3 | | | | MeiA | NM_001003811 | Yatsenko et al., 2015 |
| 3 | Patient 4 | c.1837+1G>C | p.? | 1/- | MeiA (RS [note: not validated by CREM IHC]) | NM_001003811 | Yatsenko et al., 2015 |
| 4 | Patient 5 | c.792+1G>A | p.? | 1/- | MeiA | NM_001003811 | Yatsenko et al., 2015 |
| 5 | WHT3759 | c.1259_1260insTT | p.(Trp421Cysfs*25) | 1/- | MA (pachytene) | NM_031276 | Yang et al., 2015 |
| 6 | WHT2445 | c.1793-1G>A | p.? | 1/- | azoo | NM_031276 | Yang et al., 2015 |
| 7 | 09-297 | c.(82+1_83-1)_(651+1_652_1)del | | 1/- | SPC arrest, M-I | NM_001003811.2 | Krausz et al., 2020 |
| 8 | NOA8 | c.2525G>A | p.(Trp842*) | 1/- | MA | NM_001003811.1 | Chen et al., 2020 |
| 9 | | c.151_154del | p.(Asp51Phefs*8) | 1/- | MeiA (RS [note: not validated by CREM IHC]) | NM_031276 | Yu et al., 2021 |
| 10 | P5648 | c.1796+2T>G | p.? | 1/- | MA | NM_001003811 | Ji et al., 2021 |
| 11 | P6825 | c.1426-1G>T | p.? | 1/- | MA | NM_001003811 | Ji et al., 2021 |

| | | | | | | | |
|---|---|---|---|---|---|---|---|
| 12 | P8122 | c.1253dup | p.(Asn418Lysfs*10) | 1/- | MA | NM_001003811 | Ji et al., 2021 |
| 13 | P8251 | c.298del | p.(Val100Leufs*6) | 1/- | NOA | NM_001003811 | Ji et al., 2021 |
| 14 | P5048 | c.1051G>T | p.(Glu351*) | 1/- | MA | NM_001003811 | Ji et al., 2021 |
| 15 | P9225 | c.857del | p.(Lys286Argfs*6) | 1/- | NOA | NM_001003811 | Ji et al., 2021 |
| 16 | A2799 | c.2240C>A | p.(Ser747*) | 1/- | MeiA | NM_031276 | An et al., 2021 |
| 17 | A2153 | c.1246C>T | p.(Gln416*) | 1/- | NOA | NM_031276 | An et al., 2021 |
| 18 | NOA49 | c.559_560del | p.(Met187Valfs*6) | 1/- | MA | NM_031276 | Tang et al., 2022 |
| 19 | P3 | c.313C>T | p.(Arg105*) | 1/- | MA, TESE negative | NM_031276 | Song et al., 2023 |
| 20 | M1390 | c.(159+1_160-1)_(692+1_693-1)del | | 1/- | MeiA | NM_001003811.2 | Wyrwoll et al., 2023 |

abbreviations: c = coding DNA reference sequence, p =protein reference sequence, azoo = azoospermia, crypto = cryptozoospermia, MeiA, meiotic arrest, M-I = metaphase I arrest, MA = maturation arrest, RS = round spermatid, SPC = spermatocyte, NA = no information, NOA = non-obstructive azoospermia, TESE = testicular sperm extraction

150 **Appendix Table S3. Antibody information.**

| Target | Species | Dilution, individual protocol requirements | Source |
|---|---|---|---|
| **primary antibodies** | | | |
| CREM | rabbit, polyclonal | IHC: 1:2000 pH 6 | Sigma Aldrich, #HPA001818 |
| H3S10p | rabbit, polyclonal | IHC: 1:2500, pH 9 | GeneTex, #GTX128116 |
| γH2AX | mouse, monoclonal | IHC: 1:30 in TBS + 0.01% Tween, pH 9<br>IF (meiotic spreads): 1:1000 | Merck, #05-636 |
| TEX11 | rabbit, polyclonal | IF (meiotic spreads): 1:25<br>WB: 1:500 | Sigma, #HPA002950 |
| SYCP3 | rabbit, polyclonal | IF (meiotic spreads): 1:500 | R&D Systems, #AF3750 |
| SYCP1 | rabbit, polyclonal | IF (meiotic spreads): 1:50 | Novus Biological, #NB300-228 |
| ACA | human, polyclonal | IF (meiotic spreads): 1:150 | Biozol, #15-234 |
| MLH1 | human, monoclonal | IF (meiotic spreads): 1:25 | BD Bioscience, #550838 |
| DYK | mouse, monoclonal | WB: 1:1500 | Merck, #F3165 |
| HA | rat, monoclonal | WB: 1:1500 | Sigma Aldrich, #11867423 |
| Isotype control | mouse | adapted to primary antibody | Merck, #I5381 |
| Isotype control | rabbit | adapted to primary antibody | Merck, #I5006 |
| Isotype control | goat | adapted to primary antibody | Merck, #I5256 |
| **secondary antibodies** | | | |
| anti-rabbit biotin | goat, polyclonal | IHC: 1:100 | abcam, #ab6012 |
| anti-mouse biotin | goat, polyclonal | IHC: 1:100 | abcam, #ab5886 |
| anti-mouse HRP | donkey, recombinant | WB: 1:1000 | Santa Cruz, #sc-516102 |
| anti-rat HRP | goat, polyclonal | WB: 1:1000 | Sigma-Aldrich, #A9037 |

| | | | |
|---|---|---|---|
| anti-human DyLight 405 | donkey | IF (meiotic spreads): 1:400 | Jackson Immuno, #709-475-149 |
| anti-goat Alexa Fluor 488 | donkey | IF (meiotic spreads): 1:500 | Thermo Fisher, #A11055 |
| anti-mouse Alexa Fluor 568 | donkey | IF (meiotic spreads): 1:500 | Thermo Fisher, #A10037 |
| anti-mouse Alexa Fluor Plus 647 | donkey | IF(meiotic spreads): 1:500 | Thermo Fisher, #A32787 |
| anti-rabbit Alexa Fluor 568 | donkey | IF (meiotic spreads): 1:500 | Thermo Fisher, #A10042 |
| anti-rabbit Alexa Fluor Plus 647 | donkey | IF (meiotic spreads): 1:500 | Thermo Fisher, #A32795 |
| **others** | | | |
| Streptavidin HRP | | IHC: 1:500 | Sigma-Aldrich, #S5512 |
| Fab anti-human | goat | IF (meiotic spreads): 50 µg / mL | Jackson Immuno, #109-007-003 |

**Appendix Table S4. Primer information.**

| Target | Primer sequences (5' – 3') |
|---|---|
| **Sanger sequencing** | |
| M1AP c.676dup | F: TGGGTCTGGAAATGTTGCTGA<br>R: GATTGCTAGAGCCCAGGCAT |
| M1AP c.1073_1074+10del | F: ACAGAATATATATCTAGGGCTTGACAC<br>R: GAGTCTGCTTCAACTCTTCCCA |
| SHOC1 c.1085_1086del | F: GCAGAGCCAGGGCCTATATG<br>R: AATTCAAGAGCCCCACAGCC |
| SHOC1 c.1351del + c.1347T>A | F: TGGTCCTGTGCAGTCAAGTT<br>R: ACACTCGTTTAGGTGTGGAAGT |
| SHOC1 c.945_948del | F: CCCACATTTCTACTCGTTGTACC<br>R: CTTGAATCTGGGGCGGAGG |
| SHOC1 c.1939+2T>C | F: TGTCAATTAGGAGCTTCACTGAG<br>R: TGCAGATAGCCAGTGCCAA |
| TEX11 c.450C>T | F: TGTGGAGTTCAAAGTAGAACACAGAAC<br>R: TCCAATCAGCATTAGTAACATCACC |
| TEX11 c.1425G>A | F: TGTTGACCAAGACTGATAATAAAATGC<br>R: CAGTGTGCAAATCAAGAAAATGTC |
| TEX11 c.22del | F: CGTTGCCAGGCAGACTTATG<br>R: GGGATTACCACGCCCAAC |
| TEX11 c.1096dup | F: CACTCTCCAGCACTGGATGTTAATAC<br>R: ATTGCCAAGGTTGGTCTCAAG |
| TEX11 c.792+1G>A | F: GCCAAATGGAAAAAGGCATC<br>R: ACCCAAACATTGTTCAAAGCAC |
| TEX11 c.1837+1G>C | F: ATGAGGGCACTGGGAATGAG<br>R: TCTCTGCTTGTGAATGAAGAAACC |
| TEX11 c.1245G>A | F: GGAGAATCAGGCAGCAGTACC<br>R: CAGCGATGACATTTCCCTACAC |
| TEX11 c.731G>A | F: TTCTCCAACCTGAATGTTTTGC<br>R: AAGGAAGGAAGAACACATTTTTCTATG |

| | |
|---|---|
| SPO16 Exon 4 | F: ACTACCCAGTATCTTCATGTGGA<br>R: CCACAGAGGATTTGAGATGGCT |
| pcDNA3.1 | F: GTAACAACTCCGCCCCATTG<br>R: AGGAAAGGACAGTGGGAGTG |
| SHOC1 WT (cDNA) | F: GCCAAGAATTCAAGAGCCCC<br>R: CCTGCTTGTTTCCACCACTC |
| SHOC1 WT c.287 (cDNA) | F: GTAGTAGAAAACACCTACC |
| SHOC1 WT c.803 (cDNA) | F: CTCTATTCCTAACATGCC |
| SHOC1 WT c.1324 (cDNA) | F: GCAAAAGAAGTACCAGATC |
| SHOC1 WT c.1979 (cDNA) | F: CTCTCTTACATCTTCTGG |
| SHOC1 WT c.2636 (cDNA) | F: CAGACATACTTCAGCTGC |
| SHOC1 WT c.3086 (cDNA) | F: GGTTGGATAAATCCTGGC |
| SHOC1 WT c.3746(cDNA) | F: CTCAGAAGAGAGTGTCAG |
| SHOC1 WT c.4241 (cDNA) | F: TGTGCTCACAACTACCAC |
| TEX11 WT (cDNA) | F: TGGCCTTGCGTTTCCTTAAC<br>R: ACTGGGCCCTTGTTGTTACT |
| TEX11 WT c.276 (cDNA) | F: AAGCCTCATTTGCCTCAG |
| TEX11 WT c.809 (cDNA) | F: ATAAGGCTCTCAATGCTG |
| TEX11 WT c.1404 (cDNA) | F: TGAACGACATGACCCTAG |
| TEX11 WT c.2021 (cDNA) | F: CAGTTGATCTAGAGCAAG |
| TEX11 WT c.2700 (cDNA) | F: TAGTCAGCTTGTGGAAGC |
| SPO16 WT (cDNA) | F: GTTTTGTCTGCTGCCCTCC<br>R: GCATTTACTGTGTTGTGTACTGG |
| SPO16 WT c.355 (cDNA) | F: CTTCCAGTACACAACACAG |
| **ddPCR** | |
| TEX11 WT | F: TGGGTTAACTGTGAGGAGAC<br>R: CAATTCTCCCCTCTCCCTAC |
| TEX11 (probe) | FAM-ACGCTGAGTGAAACAAGCCAGTC-BHQ1 |

| | |
|---|---|
| Reference WT (ZIC1) | F: CTCTGGCTACGAATCCTCC<br>R: CAATTCTCCCCTCTCCCTAC |
| Reference (ZIC1, probe) | HEX- CGCCTCCCACCATCGTGTCT |
| **Minigene assay** | |
| M1AP in Ex7 F-seq | F: CACCTCTGCTTCAACTCTTCCCAGT<br>R: TACACCTGGAATGCTCTGCC |
| M1AP aus Ex7 R-seq | F: TGTAAAACGACGGCCAG<br>R: AGCAGGCTGTGACACAAAGCATG |
| SHOC1 c1339+2 MiniGene | F: CACCTTCAGATAGAAGTTCGGATCTCC<br>R: TGGTTTGGCTGGGTATCACA |
| SHOC1 Ex13 | F: ACCCTCCCTACTGCTAATTGG |
| rat insulin | F: CCTGCTCATCCTCTGGGAGC<br>R: AGCAGGCTGTGACACAAAGCATG |

### Appendix Figures

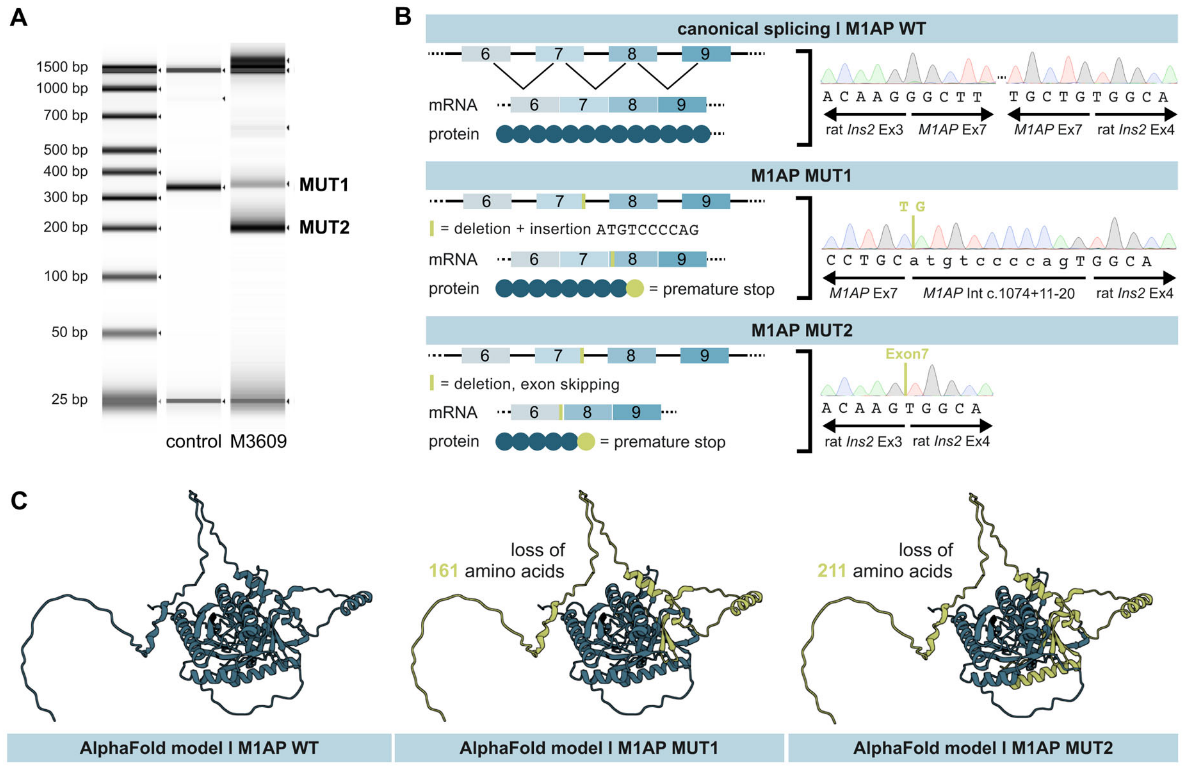


**Appendix Figure S1. *M1AP* splice site variant identified in M3609.** A. Amplified minigene cDNA encompassing – c.1073_1074+10del or respective wildtype (control/WT) sequence. B. Schematic illustration of variant effect on genomic, transcriptomic, and protein level combined with sequencing results for each minigene product reveals aberrant splicing. In the WT minigene construct, *M1AP* exon 7 (Ex7) is encompassed by two known exons of rat *Insulin 2*, exon 3 and exon 4 (rat *Ins2* Ex3/Ex4). In M3609, the variant led to two splicing products: one (MUT1) showed the recognition of a cryptic splice site leading to a frameshift and premature stop codon in *M1AP* exon 8. The second (MUT2) resulted in skipping of exon 7 and a premature stop codon in exon 8. C. Both splicing products lead to the loss of amino acids, presumably effecting M1AP's function and interaction.

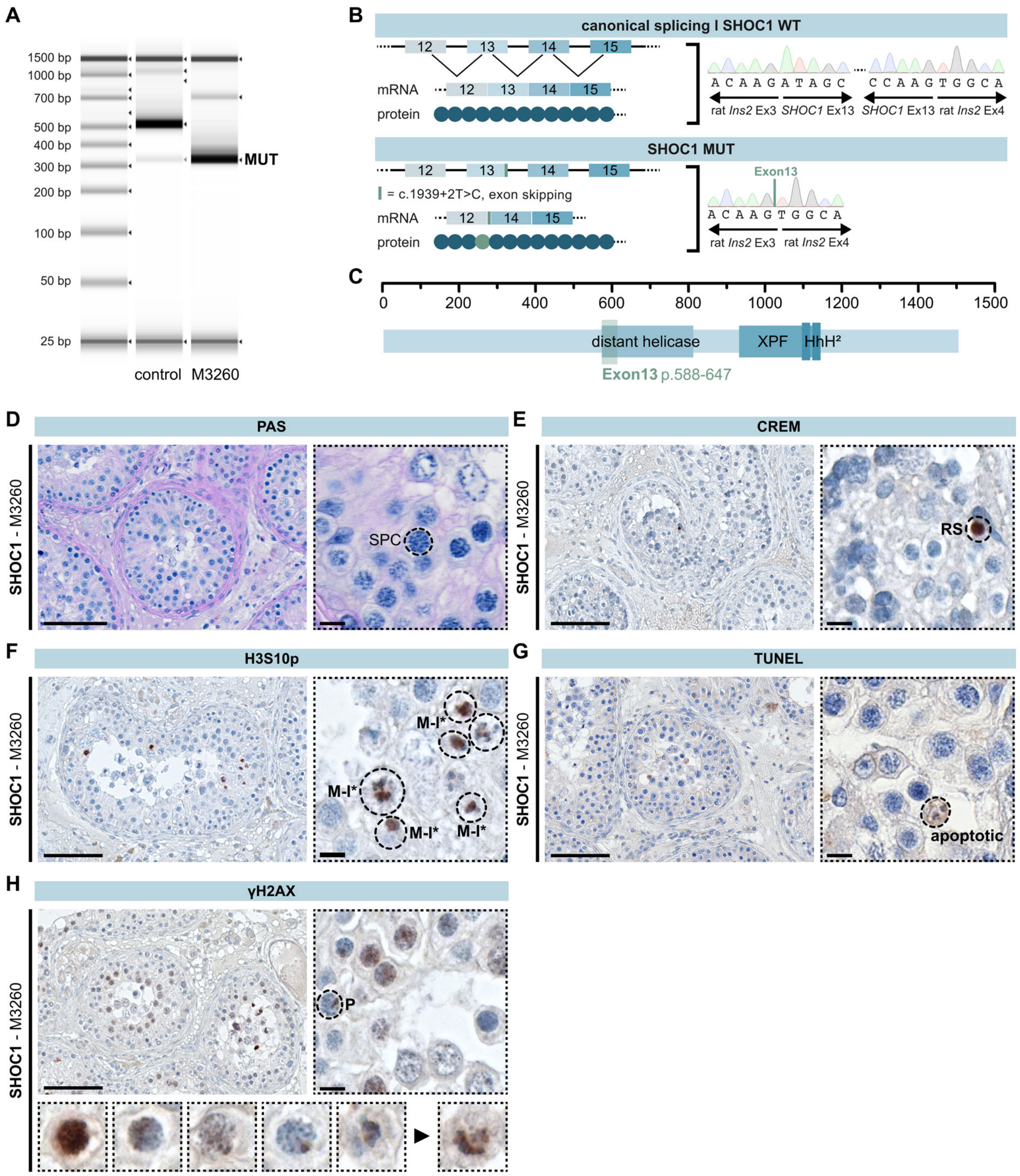


**Appendix Figure S2. *SHOC1* splice site variant identified in M3260 with predominant meiotic arrest and rare round spermatids.** A. *SHOC1* (NM_173521.5) has 26 exons and its corresponding protein comprises 1444 amino acids. Amplified minigene cDNA encompassing – c.1939+2T>C (M3260) or respective wildtype (control/WT) sequence. B. Schematic illustration of variant effect on genomic, transcriptomic, and protein level combined with sequencing results for each minigene product reveals aberrant splicing. In the WT minigene construct, *SHOC1* exon 13 (Ex13) is encompassed by two known exons of rat *Insulin* 2, exon 3 and exon 4 (rat *Ins2* Ex3/Ex4). In M3260, the variant resulted in in-frame skipping of exon 13 and a predictive loss of 59 amino acids representing 4% of the total protein. C. This affects the distant helicase hits region but not the highly conserved 'SHOC1 homology region' (amino acids 937-1105, NP_775792; Macaisne et al., 2008). This region contains an XPF endonuclease-like central and a helix-hairpin-helix (HhH$^2$) domain and is important for the XPF-ERCC1-like complex formation between SHOC1 and SPO16 (De Muyt et al., 2018; Zhang et al., 2019). Yeast studies highlighted that the N-terminal part of Zip2 is linked to the chromosome axis and the other ZMM components through Zip4 interaction, while the XPF domain interacts exclusively with Spo16 (De Muyt et al., 2018). Given that M3260 expresses all exons of *SHOC1* except for exon 13, the interaction with SPO16 and in parts with the ZMM proteins, such as TEX11, remains intact. However, a changed protein conformation due to the loss of exon 13 could influence some of these interactions and explain the observed phenotype of predominant meiotic arrest with rare round spermatids (D) that were positive for CREM-staining (E). (F) H3S10 staining showed only aberrant metaphase I-like spermatocytes (M-I*). (G) TUNEL staining showed an increased number of apoptotic spermatocytes similar to patients with complete LoF variants in *M1AP, SHOC1, TEX11,* or *SPO16.* (H) In yH2AX staining, single tubules contained pachytene-like cells (P) with a clearly distinguishable XY body were observed, which is in line with the presence of round spermatids. In addition, also aberrant pachytene-like cells were present. (I) The specific type of arrest of M3260 is described as a metaphase I arrest (MM-I) with rare round spermatids (RS).

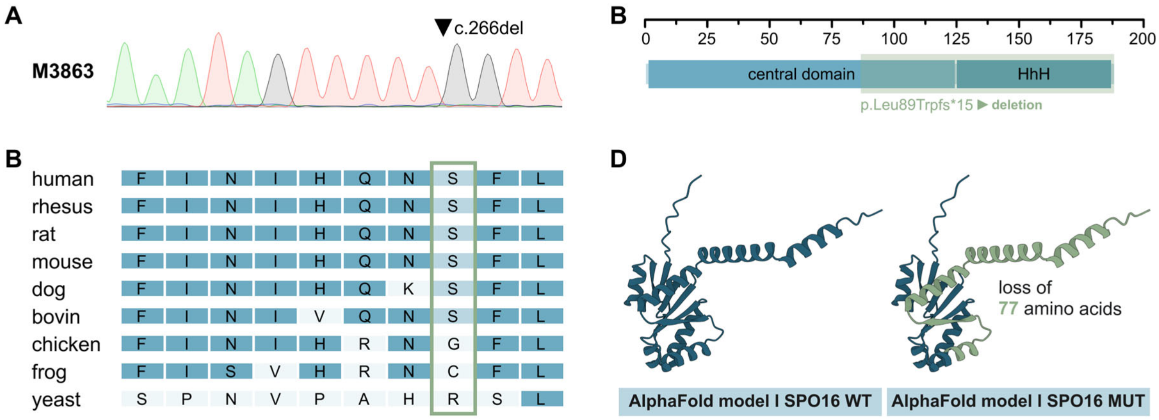


**Appendix Figure S3.** ***SPO16*** **loss-of-function variant identified in M3863.** A. Sanger sequencing of M3863 revealed the frameshift variant c.266del leading to premature stop codon (p.Leu89Trpfs*15). B. Such a truncated protein would lack the highly conserved helix-hairpin-helix (HhH$^2$) domain which is important for the XPF-ERCC1-like complex formation between SHOC1 and SPO16 (De Muyt et al., 2018; Zhang et al., 2019). C. Conversation analysis of the *SPO16* variant. D. The premature stop codon would truncate 42.5% of the complete protein.

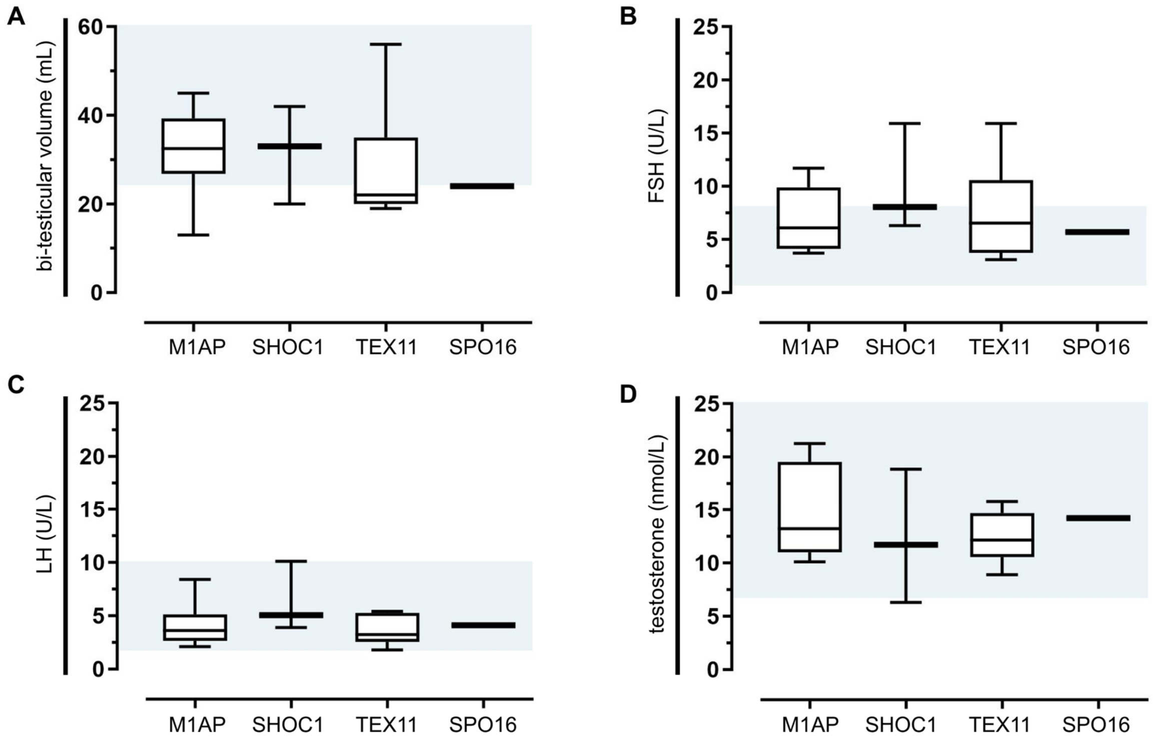


**Appendix Figure S4. Overview of clinical data of men with loss-of-function variants in *M1AP*, *SHOC1*, *TEX11* or *SPO16*.** Exome data of men with variants in *M1AP* (N=10), *SHOC1* (N=4), *TEX11* (N=9), and *SPO16* (N=1) was queried and depicted data shows the median values with the respective 95% confidence intervals. A. Bi-testicular volume (mL). B. Serum FSH (U/L). C. Serum LH (U/L). D. Serum testosterone (nmol/L). Blue areas represent respective reference values (FSH = 1-7 IU/L, LH = 2-10 IU/L, T ≥ 12 nmol/L, bi-testicular volume ≥ 24 mL per testis.)

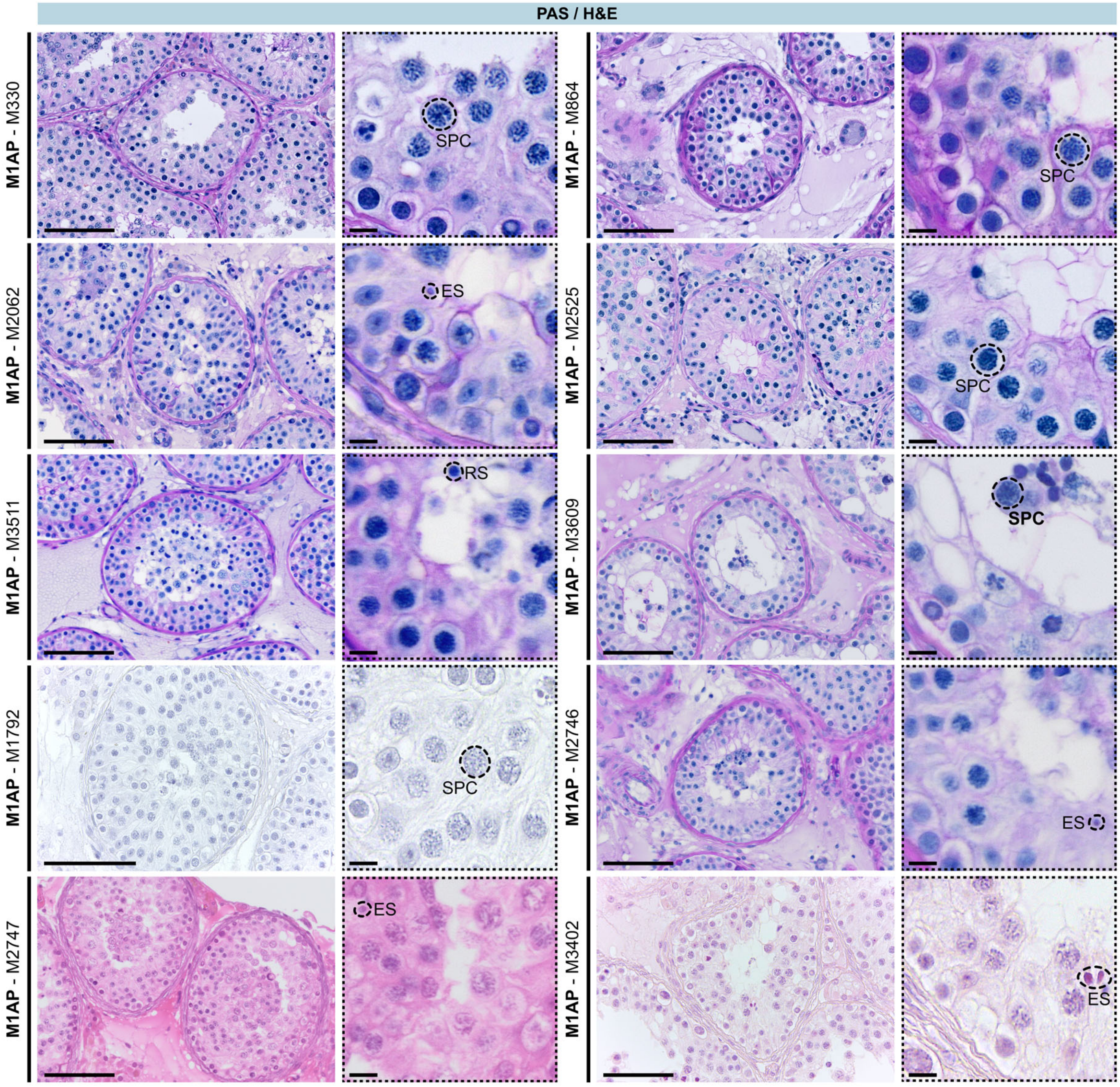


**Appendix Figure S5. PAS (N=7) or H&E (N=3) staining of men with LoF variants in *M1AP*.** All men carrying variants in *M1AP* underwent attempts for testicular sperm retrieval (TESE, N=10). Overview staining revealed testicular architecture and germ cell types were quantified for each tubule. SPC = spermatocyte, RS = round spermatid, ES = elongated spermatid. The scale bar represents 100 µm and 10 µm, respectively.

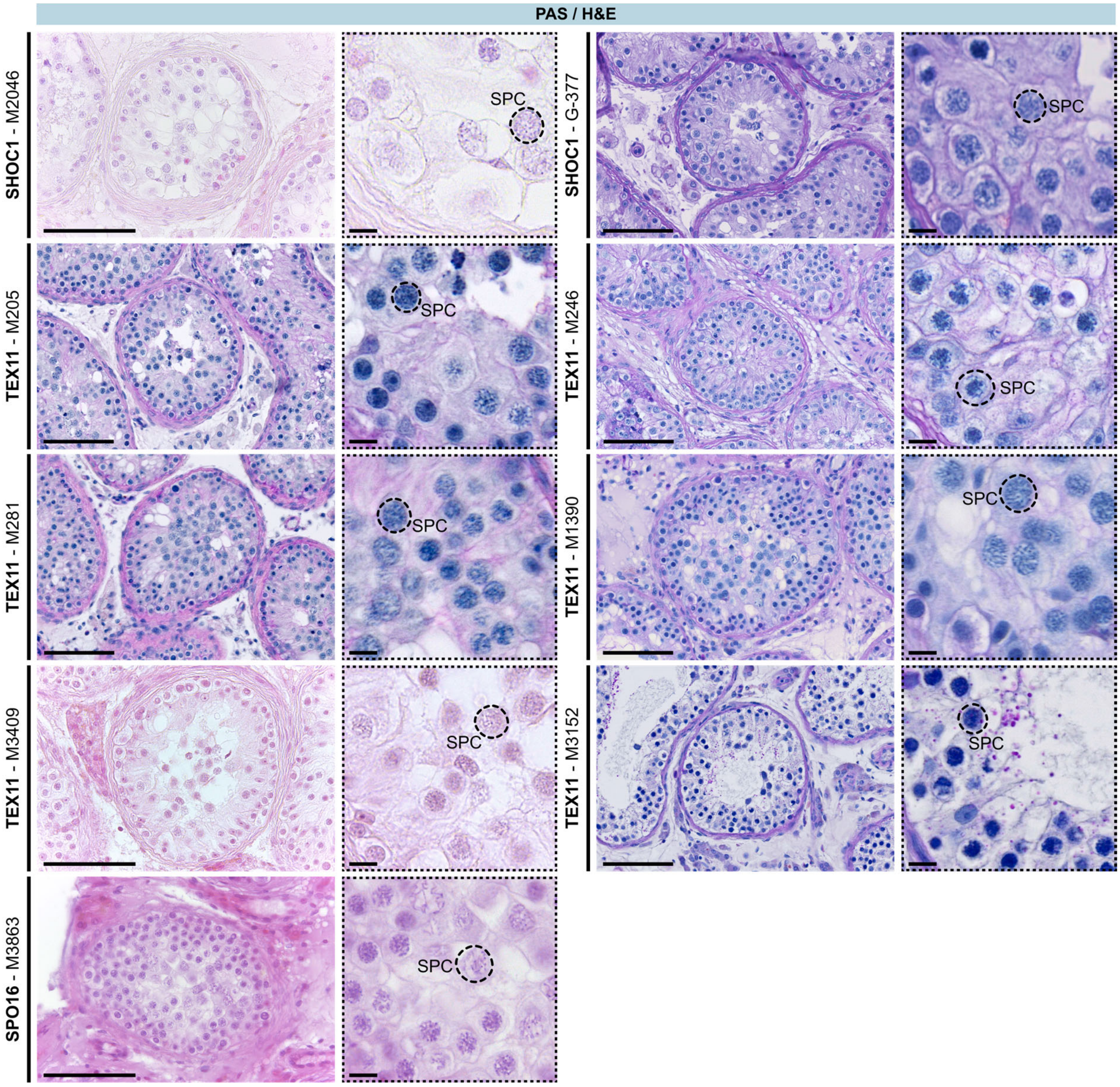


**Appendix Figure S6. PAS (N=6) or H&E (N=3) staining of men with LoF variants in *SHOC1*, *TEX11* or *SPO16*.** Ten of 14 men carrying variants in *SHOC1*, *TEX11* or *SPO16* underwent testicular surgery for TESE attempt. Overview staining revealed the testicular architecture and germ cell types were quantified for each tubule. SPC = spermatocyte. The scale bar represents 100 µm and 10 µm, respectively.

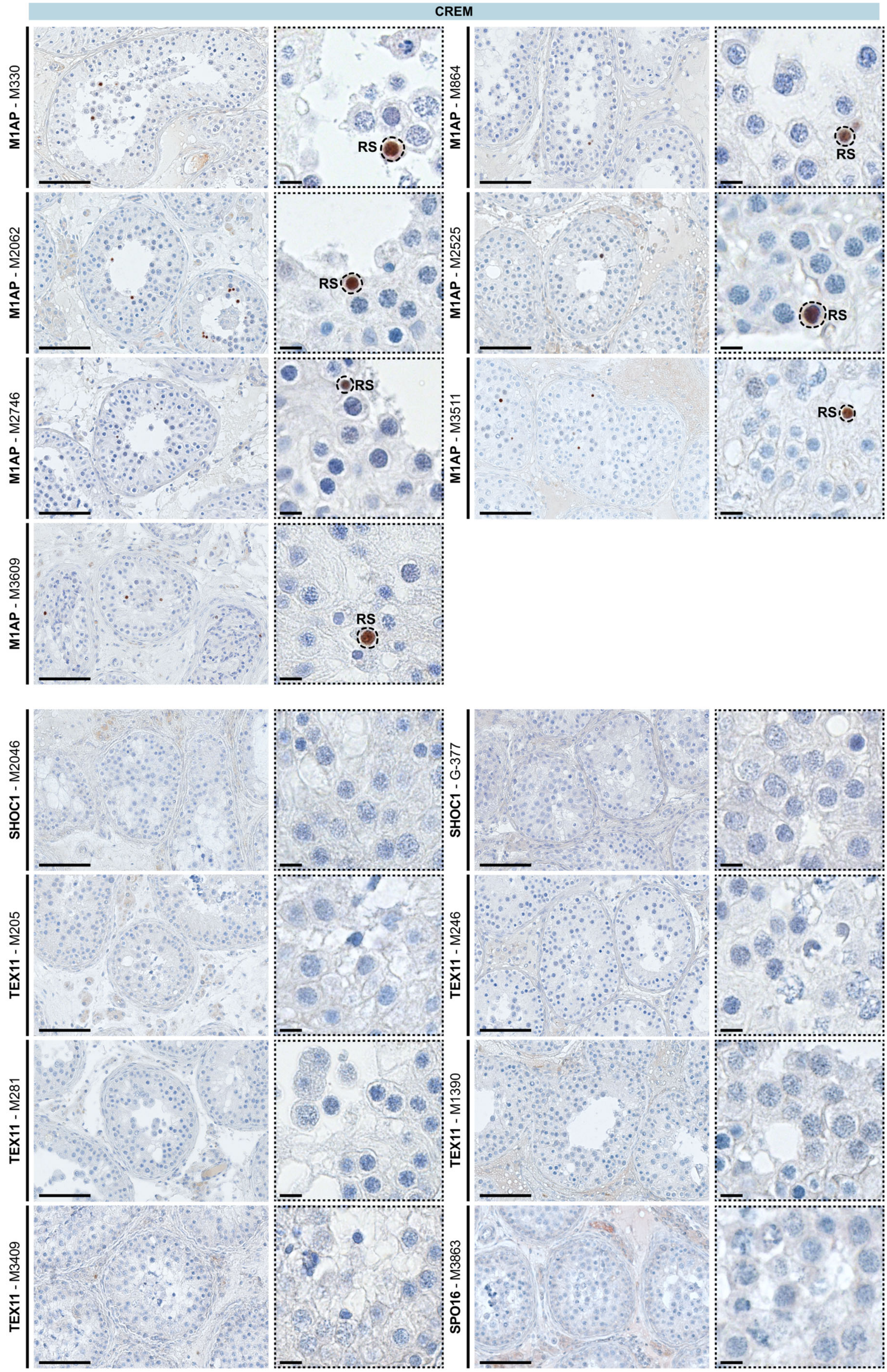
CREM
M1AP - M330
RS
M1AP - M864
RS
M1AP - M2062
RS
M1AP - M2525
RS
M1AP - M2746
RS
M1AP - M3511
RS
M1AP - M3609
RS
SHOC1 - M2046
SHOC1 - G-377
TEX11 - M205
TEX11 - M246
TEX11 - M281
TEX11 - M1390
TEX11 - M3409
SPO16 - M3863

214

**Appendix Figure S7. CREM staining in men with loss-of-function variants in *M1AP*, *SHOC1*, *TEX11* or *SPO16*.** Testicular tissue was stained for CREM to analyse development of haploid round spermatids (RS). Positive cells are indicated in the magnification. The scale bar represents 100 µm and 10 µm, respectively.

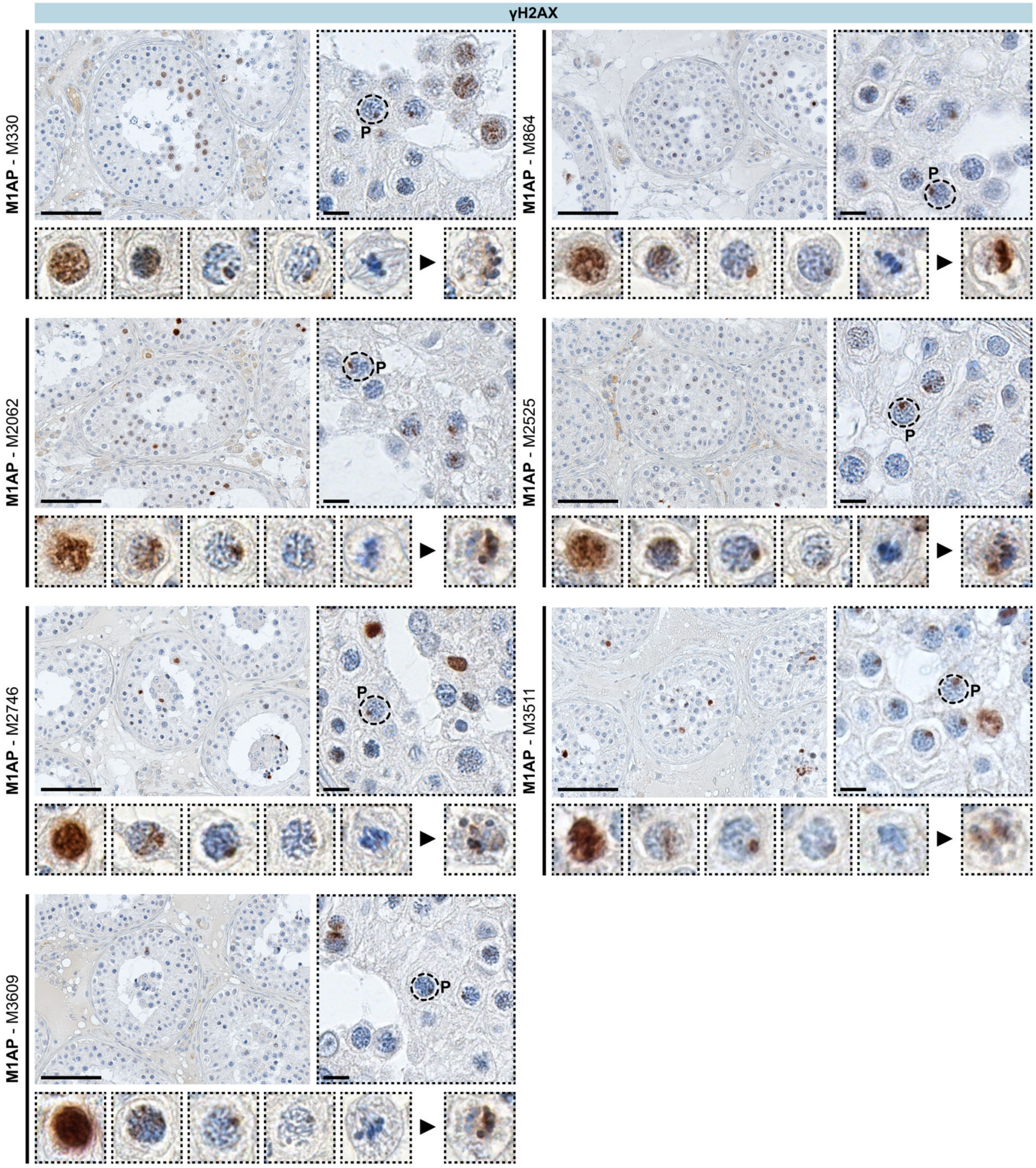


**Appendix Figure S8. γH2AX localisation showed meiosis prophase I progression in men with loss-of-function variants in *M1AP*.** Testicular tissue was stained for the DSB marker γH2AX. Meiotic prophase I substages (L = leptotene-, Z = zygotene-, P = pachytene-, D = diplotene-like) and metaphase I (M-I)-like were identified and depicted in the detail view for each man. Besides, aberrant, γH2AX-positive metaphase-like cells are shown (black arrow head). Pachytene-like cells are indicated in the magnification. The scale bar represents 100 µm and 10 µm, respectively.

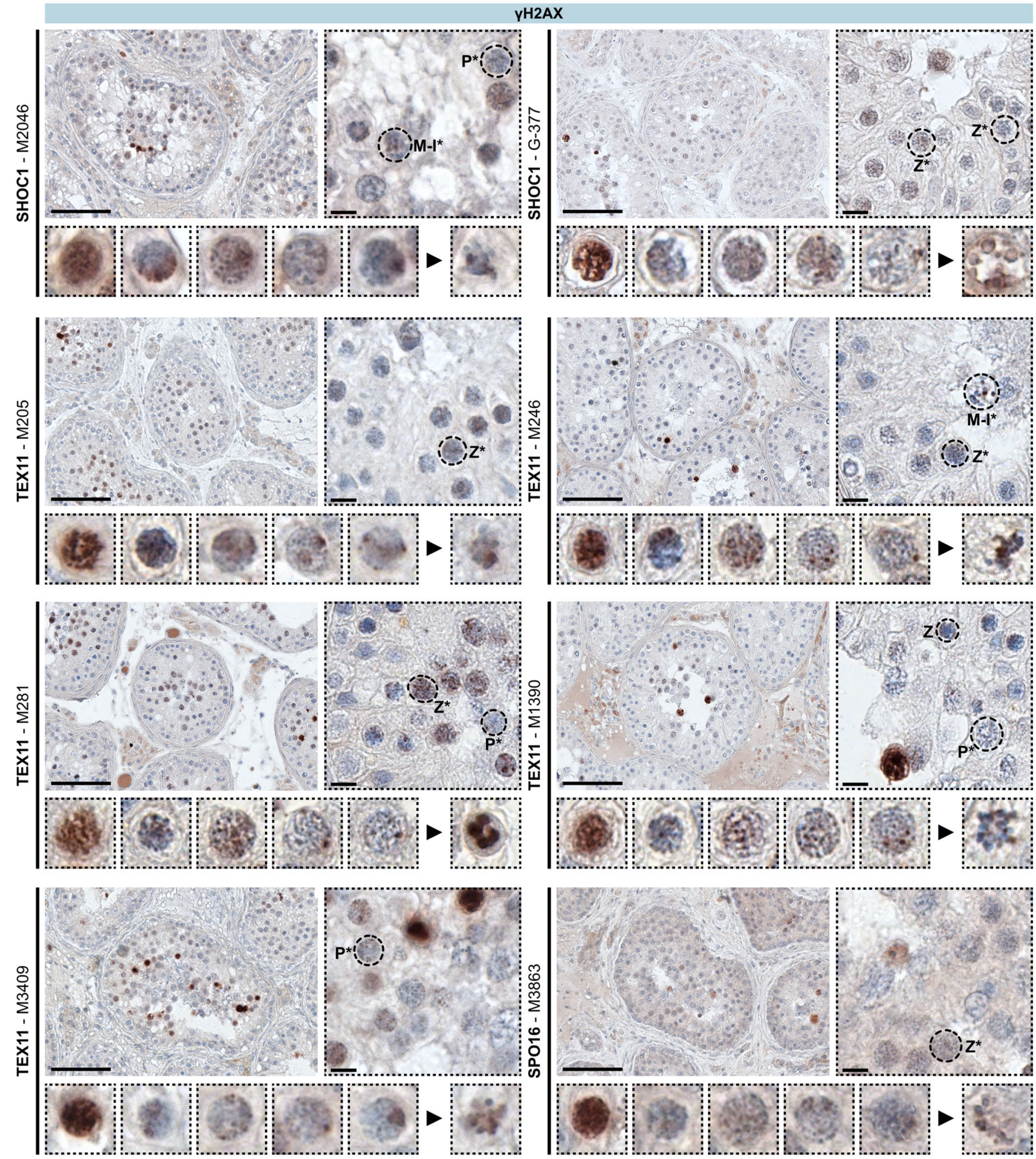


**Appendix Figure S9. γH2AX localisation showed impaired meiosis prophase I progression in men with loss-of-function variants in *SHOC1*, *TEX11* or *SPO16*.** Testicular tissue was stained for the DSB marker γH2AX. In M2046 and G-377, the majority of cells reached only a zygotene-like stage. Contrary to the other men with LoF variants in *TEX11*, one man with a frameshift variant in *TEX11* (M3409) had few pachytene-like cells in two of 86 counted seminiferous tubules. These cells had an enlarged XY body-like structure with accumulated γH2AX. Meiotic prophase I substages (L = leptotene-, Z = zygotene-like) and arrested zygotene- (Z*), pachytene- (P*) and metaphase I-like cells (M-I*) were identified and depicted in a detail view. The scale bar represents 100 µm and 10 µm, respectively.

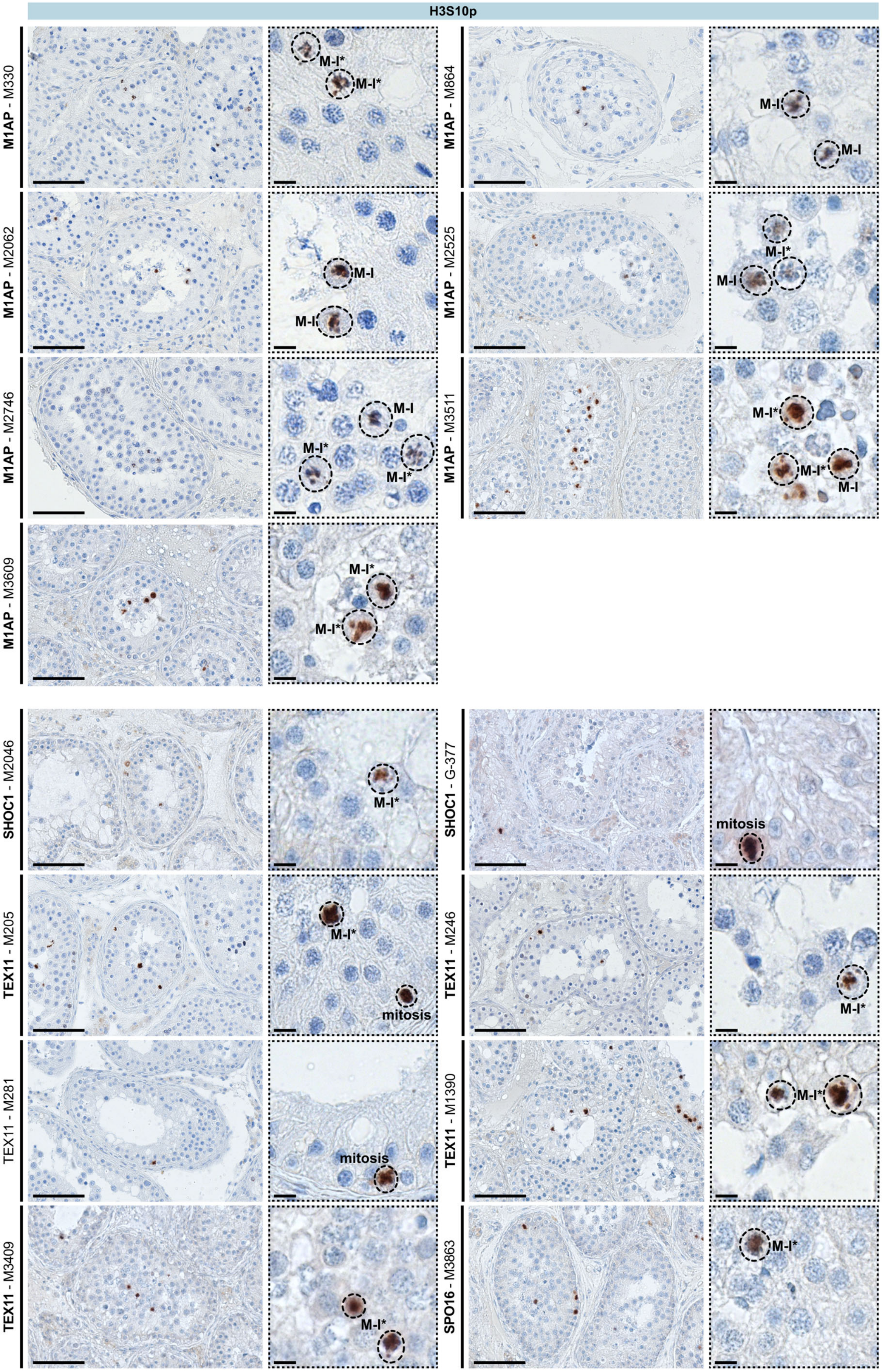


**Appendix Figure S10. Metaphase I cells were determined by H3S10p localisation in men with loss-of-function variants in *M1AP*, *SHOC1*, *TEX11* or *SPO16*.** Testicular tissue was stained for H3S10p to detect diakinesis / aberrant metaphase-I-like (M-I*) or normal metaphase I-like (M-I) cells. Localisation of positive cells was taken into account to distinguish between mitotic (spermatogonia = mitosis) and meiotic (spermatocyte) M-I cells (indicated in magnification). The scale bar represents 100 µm and 10 µm, respectively.

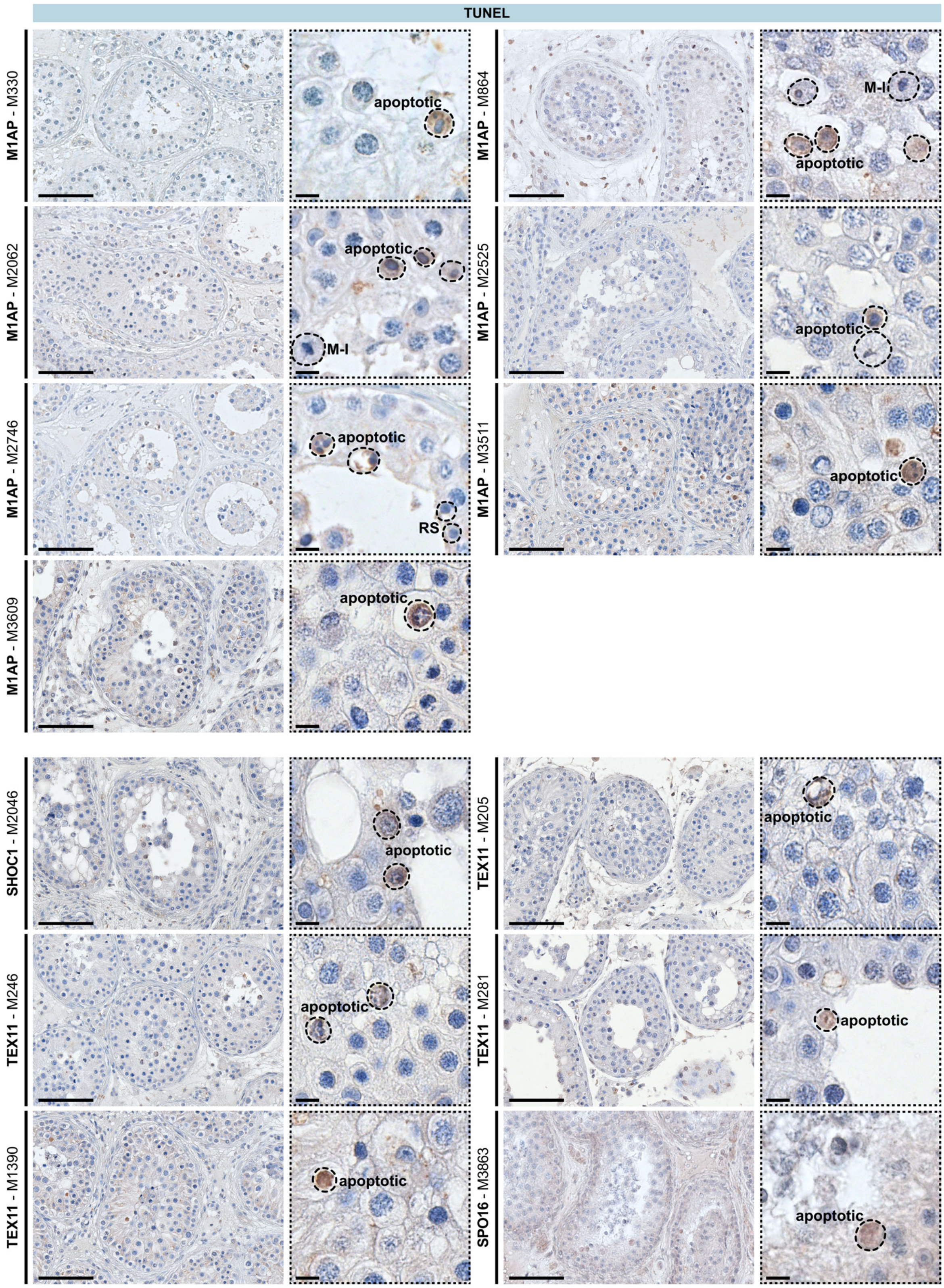


**Appendix Figure S11. Quantification of apoptosis in men with loss-of-function variants in *M1AP*, *SHOC1*, *TEX11* or *SPO16* via TUNEL.** Testicular tissue was analysed by TUNEL assay to show apoptotic cells. Positive apoptotic cells, negative diakinesis / metaphase-I-like(M-I) cells, and negative round spermatids (RS) are indicated in the magnification. The scale bar represents 100 µm and 10 µm, respectively.

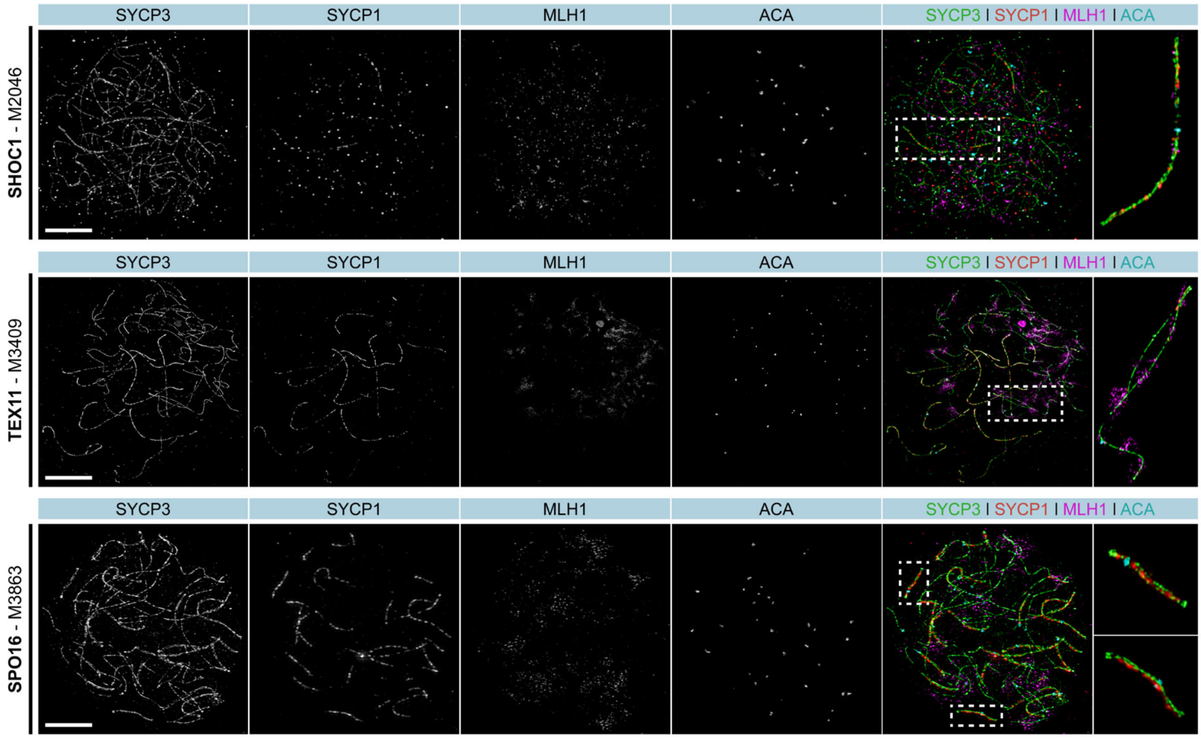


**Appendix Figure S12. Human spermatocyte spreads showed the absence of crossover in ZZS cases.** A. Meiosis-specific marker for SC assembly (SYCP3 = green), chromosome synapsis (SYCP1 = red), and crossover resolution (MLH1 = magenta) were stained on spermatocytes from representative men with loss-of-function (LoF) variants in one of the ZZS genes (*SHOC1* = M2046, *TEX11* = M3409, and *SPO16* = M3863). ACA (cyan) was used to distinguish homologous chromosomes. LoF in *SHOC1*, *TEX11*, and *SPO16* led to asynapsed chromosomes and an early meiotic arrest, where the pachytene stage was never reached and crossover events (MLH1 foci) were completely absent on chromosome axes. The scale bar represents 10 µm.

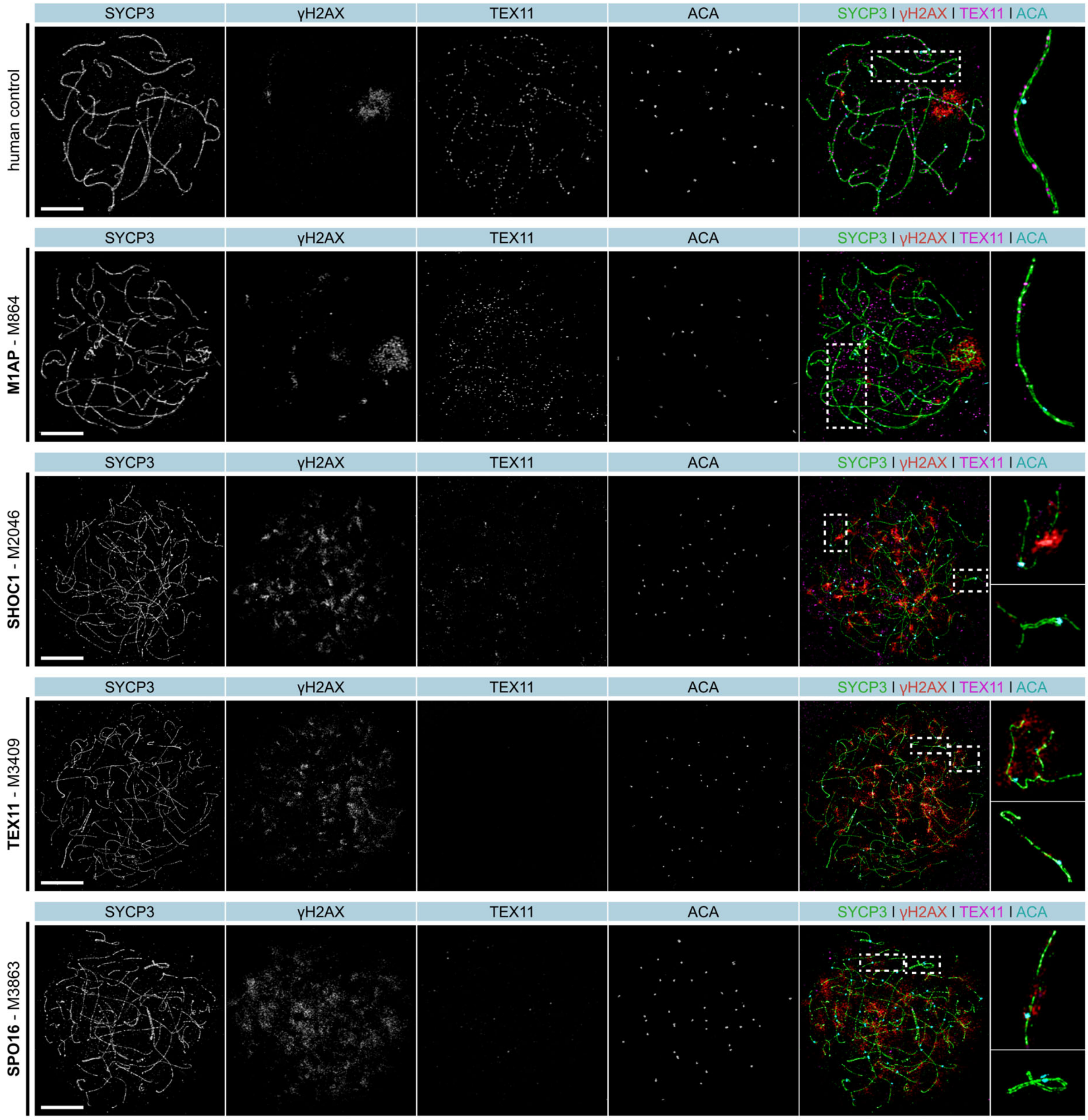


**Appendix Figure S13. Human spermatocyte spreads showed meiotic delay in ZZS cases.** Meiosis-specific marker for synaptonemal complex assembly (SYCP3 = green), DNA double-strand breaks (γH2AX = red), and ZZS recruitment (TEX11 = magenta) were stained on spermatocytes from representative men with loss-of-function (LoF) variants in *M1AP* or one of the ZZS genes (*M1AP* = M864, *SHOC1* = M2046, *TEX11* = M3409, and *SPO16* = M3863). ACA (cyan) was used to distinguish homologous chromosomes. LoF in *SHOC1*, *TEX11*, and *SPO16* was associated with an early meiosis I defect including incomplete DSB repair and disturbed assembly of early recombination intermediates (TEX11 foci), whereas the man with a LoF variant in *M1AP* had qualitatively normal meiosis I progression. However, the recruitment of TEX11 to the chromosomal axis was reduced compared to a control with qualitatively and quantitatively normal spermatogenesis. The scale bar represents 10 µm.

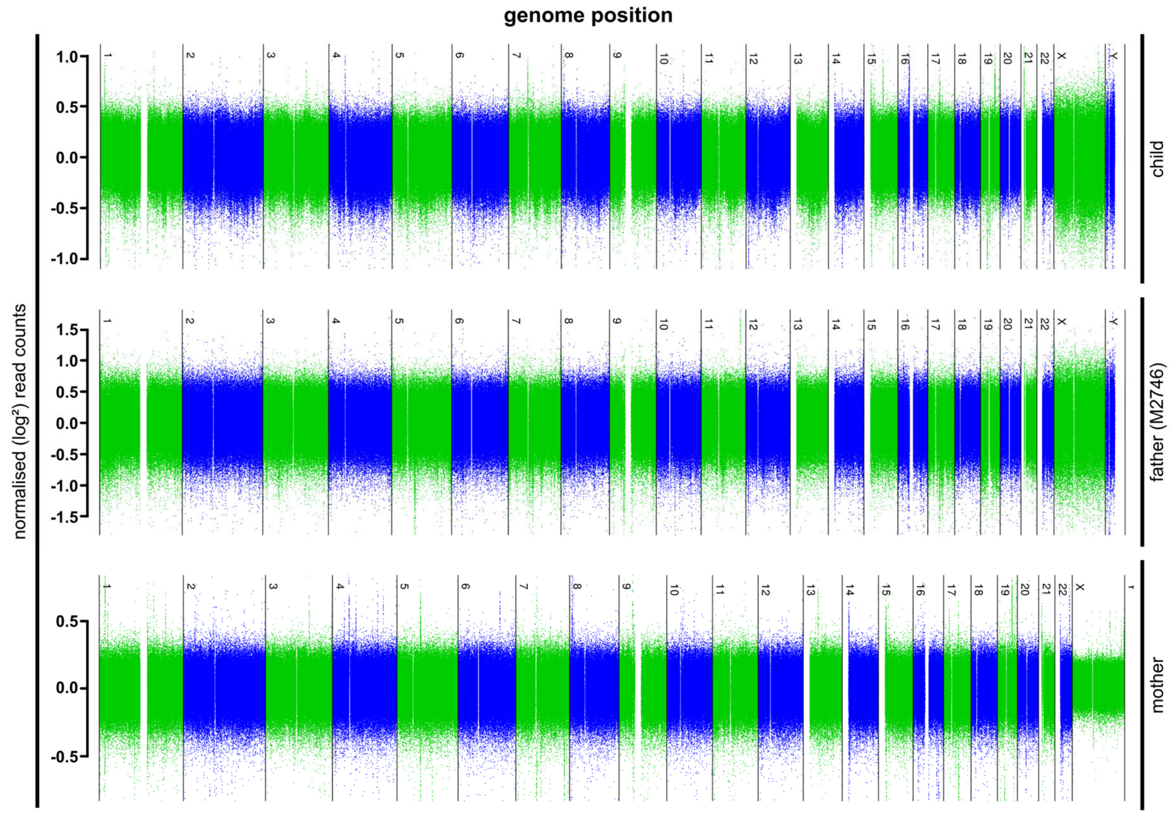


**Appendix Figure S14. Euploidy analysis of M2746, his child, and the child's mother.** Genome sequencing data was queried and read counts were normalised by dividing the median read count of each chromosome by the median read count of all autosomes. Normalised (log$^2$) read counts of autosomes (0.97 to 1.06) and of gonosomes (0.49 to 0.51) gave no evidences for chromosome aneuploidies.

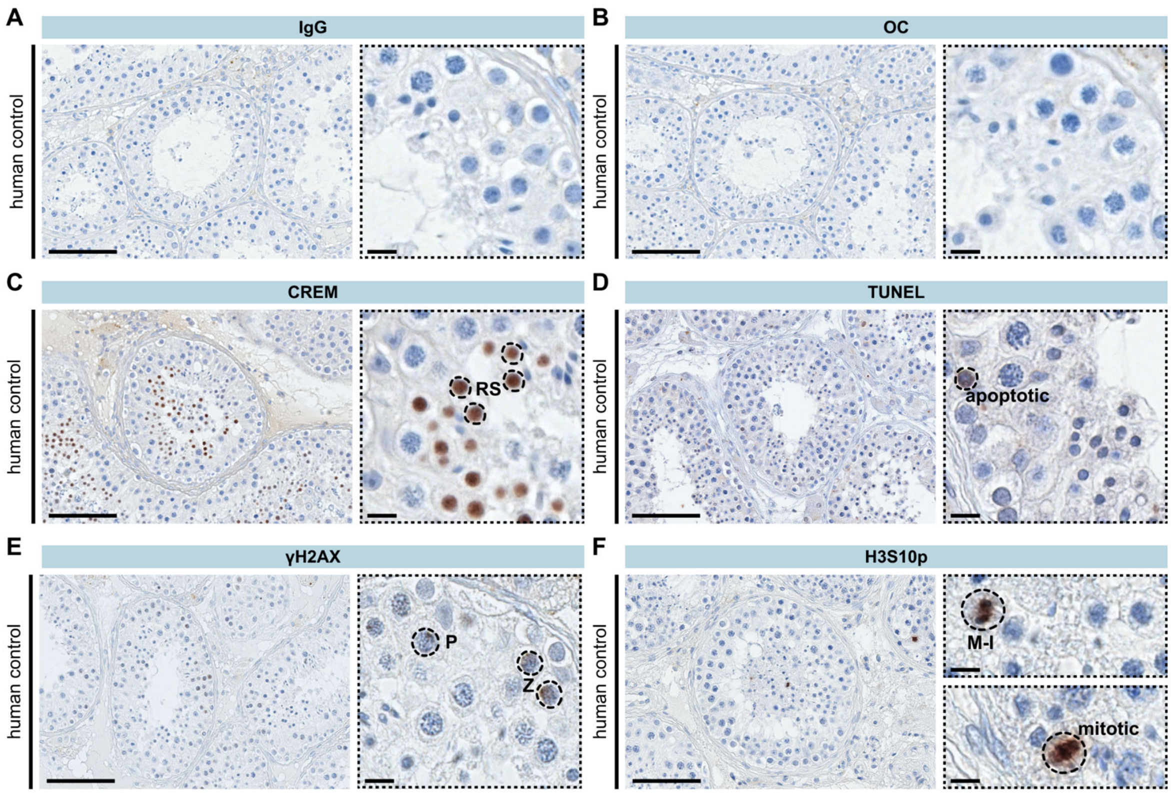


**Appendix Figure S15. Staining of testicular tissue from a representative human control.** A. Isoptype control (IgG). B. Omission of first antibody control (OC). C. CREM staining. D. TUNEL assay. E. γH2AX staining. F. H3S10p staining. Positive cells are indicated in the magnification. The scale bar represents 100 µm and 10 µm.

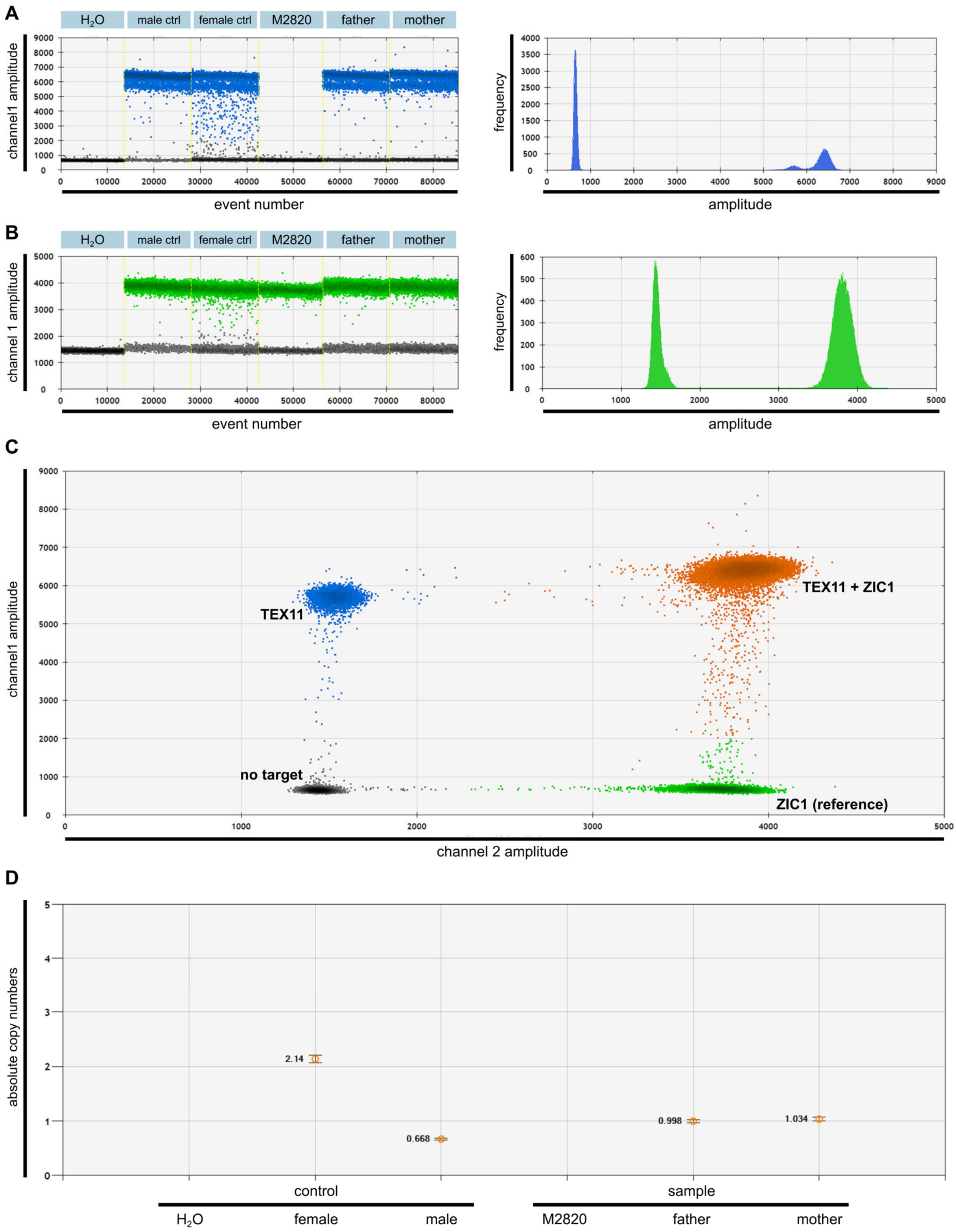


**Appendix Figure S16. Digital droplet PCR confirmed the deletion of *TEX11* exons 1-11 in M2820.** A./B. The 1D-plot showed a clear division of positive droplets (blue and green bands/peaks) and negative droplets (grey bands/peaks), which is an important quality parameter. Depicted are the *TEX11* (A) and a reference (ZIC1, B) measurements. C. The 2D-plot with four distinct droplet groups (6-FAM-positive droplets (blue), HEX-positive droplets (green), double positive droplets (orange) and negative droplets (grey)) confirmed the probe specificity. D. Calculated copy numbers showed the deletion of the respective region of *TEX11* in M2820. The $H_2O$ control did not show any fluorescent droplets, as expected. The female control shows a copy number of two and the male control shows a copy number of one, as *TEX11* is a X-chromosomal gene. M2820's father carries one copy, as expected. However, his mother only shows one copy and therefore is a heterozygous carrier of the deletion.

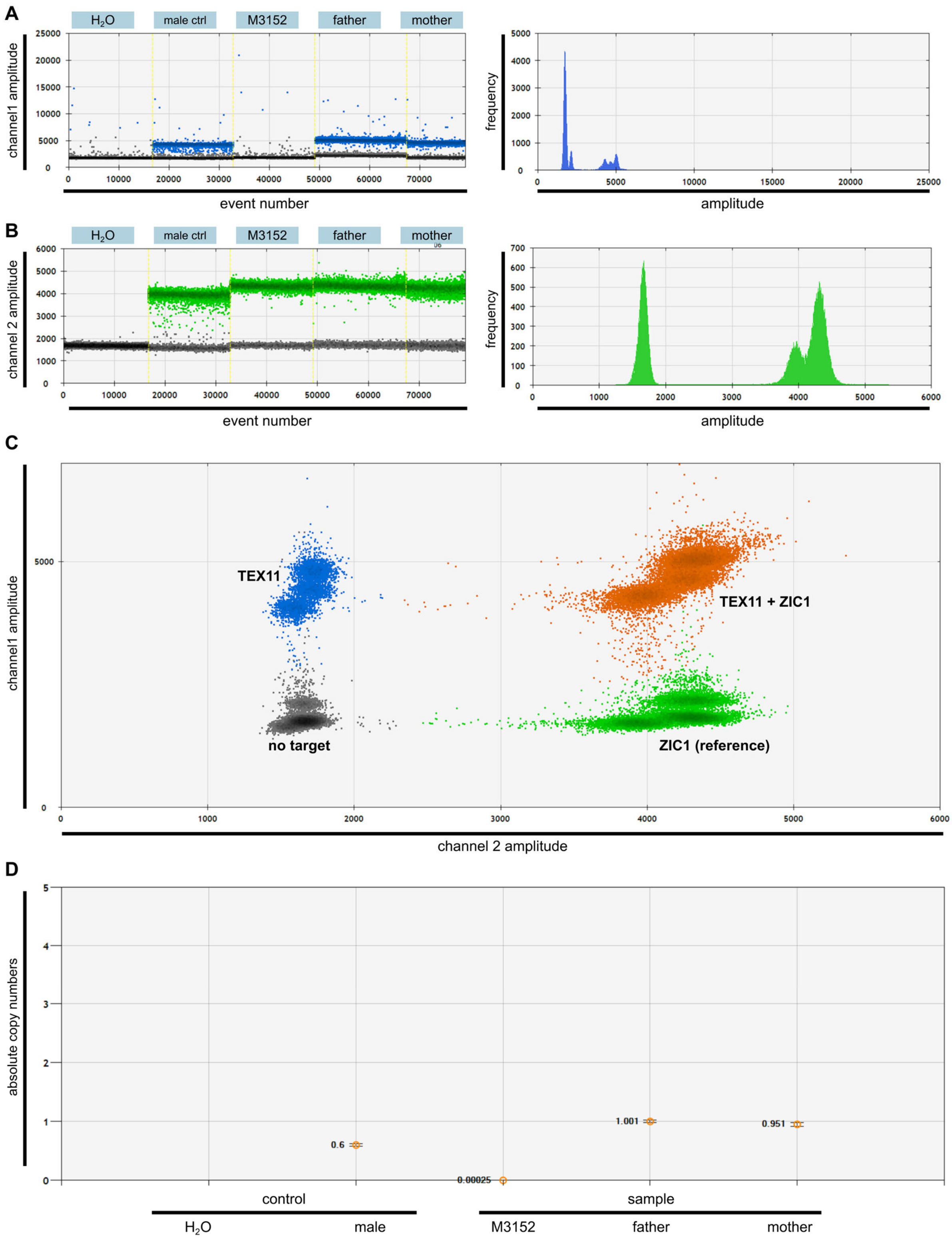


**Appendix Figure S17. Digital droplet PCR confirms the deletion of *TEX11* exons 10-11 in M3152.** A./B. Positive and negative droplets were clearly distinguishable in the *TEX11* and reference measurement (ZIC1). C. Quality parameter confirmed probe specificity. D. Calculated copy numbers showed the deletion of the respective region of *TEX11* in M3152. The $H_2O$ control did not show any fluorescent droplets and the male control showed a copy number of one, as expected for a X-chromosomal gene. M3152's father carried one copy, as expected. However, his mother only showed one copy and therefore is a heterozygous carrier of the deletion.

Karczewski, K.J., Francioli, L.C., Tiao, G., Cummings, B.B., Alföldi, J., Wang, Q., Collins, R.L., Laricchia, K.M., Ganna, A., Birnbaum, D.P., Gauthier, L.D., Brand, H., Solomonson, M., Watts, N.A., Rhodes, D., Singer-Berk, M., England, E.M., Seaby, E.G., Kosmicki, J.A., Walters, R.K., Tashman, K., Farjoun, Y., Banks, E., Poterba, T., Wang, A., Seed, C., Whiffin, N., Chong, J.X., Samocha, K.E., Pierce-Hoffman, E., Zappala, Z., O'Donnell-Luria, A.H., Minikel, E.V., Weisburd, B., Lek, M., Ware, J.S., Vittal, C., Armean, I.M., Bergelson, L., Cibulskis, K., Connolly, K.M., Covarrubias, M., Donnelly, S., Ferriera, S., Gabriel, S., Gentry, J., Gupta, N., Jeandet, T., Kaplan, D., Llanwarne, C., Munshi, R., Novod, S., Petrillo, N., Roazen, D., Ruano-Rubio, V., Saltzman, A., Schleicher, M., Soto, J., Tibbetts, K., Tolonen, C., Wade, G., Talkowski, M.E., Aguilar Salinas, C.A., Ahmad, T., Albert, C.M., Ardissino, D., Atzmon, G., Barnard, J., Beaugerie, L., Benjamin, E.J., Boehnke, M., Bonnycastle, L.L., Bottinger, E.P., Bowden, D.W., Bown, M.J., Chambers, J.C., Chan, J.C., Chasman, D., Cho, J., Chung, M.K., Cohen, B., Correa, A., Dabelea, D., Daly, M.J., Darbar, D., Duggirala, R., Dupuis, J., Ellinor, P.T., Elosua, R., Erdmann, J., Esko, T., Färkkilä, M., Florez, J., Franke, A., Getz, G., Glaser, B., Glatt, S.J., Goldstein, D., Gonzalez, C., Groop, L., Haiman, C., Hanis, C., Harms, M., Hiltunen, M., Holi, M.M., Hultman, C.M., Kallela, M., Kaprio, J., Kathiresan, S., Kim, B.J., Kim, Y.J., Kirov, G., Kooner, J., Koskinen, S., Krumholz, H.M., Kugathasan, S., Kwak, S.H., Laakso, M., Lehtimäki, T., Loos, R.J.F., Lubitz, S.A., Ma, R.C.W., MacArthur, D.G., Marrugat, J., Mattila, K.M., McCarroll, S., McCarthy, M.I., McGovern, D., McPherson, R., Meigs, J.B., Melander, O., Metspalu, A., Neale, B.M., Nilsson, P.M., O'Donovan, M.C., Ongur, D., Orozco, L., Owen, M.J., Palmer, C.N.A., Palotie, A., Park, K.S., Pato, C., Pulver, A.E., Rahman, N., Remes, A.M., Rioux, J.D., Ripatti, S., Roden, D.M., Saleheen, D., Salomaa, V., Samani, N.J., Scharf, J., Schunkert, H., Shoemaker, M.B., Sklar, P., Soininen, H., Sokol, H., Spector, T., Sullivan, P.F., Suvisaari, J., Tai, E.S., Teo, Y.Y., Tiinamaija, T., Tsuang, M., Turner, D., Tusie-Luna, T., Vartiainen, E., Watkins, H., Weersma, R.K., Wessman, M., Wilson, J.G., Xavier, R.J., 2020. The mutational constraint spectrum quantified from variation in 141,456 humans. Nature 581, 434–443.

Khan, M.R., Akbari, A., Nicholas, T.J., Castillo-Madeen, H., Ajmal, M., Haq, T.U., Laan, M., Quinlan, A.R., Ahuja, J.S., Shah, A.A., Conrad, D.F., 2023. Genome sequencing of Pakistani families with male infertility identifies deleterious genotypes in SPAG6, CCDC9, TKTL1, TUBA3C, and M1AP. Andrology 1–12.

Kishore, S., Khanna, A., Stamm, S., 2008. Rapid generation of splicing reporters with pSpliceExpress. Gene 427, 104–110.

Krausz, C., Riera-Escamilla, A., Moreno-Mendoza, D., Holleman, K., Cioppi, F., Algaba, F., Pybus, M., Friedrich, C., Wyrwoll, M.J., Casamonti, E., Pietroforte, S., Nagirnaja, L., Lopes, A.M., Kliesch, S., Pilatz, A., Carrell, D.T., Conrad, D.F., Ars, E., Ruiz-Castañé, E., Aston, K.I., Baarends, W.M., Tüttelmann, F., 2020. Genetic dissection of spermatogenic arrest through exome analysis: clinical implications for the management of azoospermic men. Genet. Med. 22, 1956–1966.

Li, H., Durbin, R., 2010. Fast and accurate long-read alignment with Burrows-Wheeler transform. Bioinformatics 26, 589–595.

Li, Y., Wu, Y., Khan, I., Zhou, J., Lu, Y., Ye, J., Liu, J., Xie, X., Hu, C., Jiang, H., Fan, S., Zhang, H., Zhang, Y., Jiang, X., Xu, B., Ma, H., Shi, Q., 2023. M1AP interacts with the mammalian ZZS complex and promotes male meiotic recombination. EMBO Rep. 24, e55778.

Macaisne, N., Novatchkova, M., Peirera, L., Vezon, D., Jolivet, S., Froger, N., Chelysheva, L., Grelon, M., Mercier, R., 2008. SHOC1, an XPF Endonuclease-Related Protein, Is Essential for the Formation of Class I Meiotic Crossovers. Curr. Biol. 18, 1432–1437.

Martin, M., 2011. Cutadapt removes adapter sequences from high-throughput sequencing reads. EMBnet.journal 17, 10.

McKenna, A., Hanna, M., Banks, E., Sivachenko, A., Cibulskis, K., Kernytsky, A., Garimella, K., Altshuler, D., Gabriel, S., Daly, M., DePristo, M.A., 2010. The genome analysis toolkit: A MapReduce framework for analyzing next-generation DNA sequencing data. Genome Res. 20, 1297–1303.

McLaren, W., Gil, L., Hunt, S.E., Riat, H.S., Ritchie, G.R.S., Thormann, A., Flicek, P., Cunningham, F., 2016. The Ensembl Variant Effect Predictor. Genome Biol. 17, 122.

Nagirnaja, L., Lopes, A.M., Charng, W.L., Miller, B., Stakaitis, R., Golubickaite, I., Stendahl, A., Luan, T., Friedrich, C., Mahyari, E., Fadial, E., Kasak, L., Vigh-Conrad, K., Oud, M.S., Xavier, M.J., Cheers, S.R., James, E.R., Guo, J., Jenkins, T.G., Riera-Escamilla, A., Barros, A., Carvalho, F., Fernandes, S., Gonçalves, J., Gurnett, C.A., Jørgensen, N., Jezek, D., Jungheim, E.S., Kliesch, S., McLachlan, R.I., Omurtag, K.R., Pilatz, A., Sandlow, J.I., Smith, J., Eisenberg, M.L., Hotaling, J.M., Jarvi, K.A., Punab, M., Rajpert-De Meyts, E., Carrell, D.T., Krausz, C., Laan, M., O'Bryan, M.K., Schlegel, P.N., Tüttelmann, F., Veltman, J.A., Almstrup, K., Aston, K.I., Conrad, D.F., 2022. Diverse monogenic subforms of human spermatogenic failure. Nat. Commun. 13, 7953.

Song, J., Sha, Y., Liu, X., Zeng, X., 2023. Novel mutations of TEX11 are associated with non-obstructive azoospermia 1–8.

Tang, D., Li, K., Geng, H., Xu, C., Lv, M., Gao, Y., Wang, G., Yu, H., Shao, Z., Shen, Q., Jiang, H., Zhang, X., He, X., Cao, Y., 2022. Identification of deleterious variants in patients with male infertility due to idiopathic non-obstructive azoospermia. Reprod. Biol. Endocrinol. 20, 1–11.

Tu, C., Wang, Y., Nie, H., Meng, L., Wang, W., Li, Y., Li, D., Zhang, H., Lu, G., Lin, G., Tan, Y.Q., Du, J., 2020. An M1AP homozygous splice-site mutation associated with severe oligozoospermia in a consanguineous family. Clin. Genet. 97, 741–746.

Varadi, M., Bertoni, D., Magana, P., Paramval, U., Pidruchna, I., Radhakrishnan, M., Tsenkov, M., Nair, S., Mirdita, M., Yeo, J., Kovalevskiy, O., Tuny, K., Žídek, A., Tomlinson, H., Hariharan, D., Abrahamson, J., Green, T., Jumper, J., Hassabis, D., Velankar, S., 2024. AlphaF old Prot ein Structure D atabase in 2024: 1–8.

Wang, W., Meng, L., He, J., Su, L., Li, Y., Tan, C., Xu, X., Nie, H., Zhang, H., Du, J., Lu, G., Luo, M., Lin, G., Tu, C., Tan, Y.Q., 2022. Bi-Allelic variants in SHOC1 cause non-obstructive azoospermia with meiosis arrest in humans and mice. Mol. Hum. Reprod. 28, 1–13.

Wyrwoll, M.J., Köckerling, N., Vockel, M., Dicke, A.K., Rotte, N., Pohl, E., Emich, J., Wöste, M., Ruckert, C., Wabschke, R., Seggewiss, J., Ledig, S., Tewes, A.C., Stratis, Y., Cremers, J.F., Wistuba, J., Krallmann, C., Kliesch, S., Röpke, A., Stallmeyer, B., Friedrich, C., Tüttelmann, F., 2023. Genetic Architecture of Azoospermia—Time to Advance the Standard of Care. Eur. Urol. 83, 452–462.

Wyrwoll, M.J., Temel, Ş.G., Nagirnaja, L., Oud, M.S., Lopes, A.M., van der Heijden, G.W., Heald, J.S., Rotte, N., Wistuba, J., Wöste, M., Ledig, S., Krenz, H., Smits, R.M., Carvalho, F., Gonçalves, J., Fietz, D., Türkgenç, B., Ergören, M.C., Çetinkaya, M., Başar, M., Kahraman, S., McEleny, K., Xavier, M.J., Turner, H., Pilatz, A., Röpke, A., Dugas, M., Kliesch, S., Neuhaus, N., Aston, K.I., Conrad, D.F., Veltman, J.A., Friedrich, C., Tüttelmann, F., 2020. Bi-allelic Mutations in M1AP Are a Frequent Cause of Meiotic Arrest and Severely Impaired Spermatogenesis Leading to Male Infertility. Am. J. Hum. Genet. 107, 342–351.

Yang, F., Silber, S., Leu, N.A., Oates, R.D., Marszalek, J.D., Skaletsky, H., Brown, L.G., Rozen, S., Page, D.C., Wang, P.J., 2015. TEX11 is mutated in infertile men with azoospermia and regulates genome-wide recombination rates in mouse. EMBO Mol. Med. 7, 1198–1210.

Yao, C., Yang, C., Zhao, L., Li, P., Tian, R., Chen, H., Guo, Y., Huang, Y., Zhi, E., Zhai, J., Sun, H., Zhang, J., Hong, Y., Zhang, L., Ji, Z., Zhang, F., Zhou, Z., Li, Z., 2021. Bi-allelic SHOC1 loss-of-function mutations cause meiotic arrest and non-obstructive azoospermia. J. Med. Genet. 58, 679–686.

Yatsenko, A.N., Georgiadis, A.P., Röpke, A., Berman, A.J., Jaffe, T., Olszewska, M., Westernströer, B., Sanfilippo, J., Kurpisz, M., Rajkovic, A., Yatsenko, S.A., Kliesch, S., Schlatt, S., Tüttelmann, F., 2015. X-Linked TEX11 Mutations, Meiotic Arrest, and Azoospermia in Infertile Men. N. Engl. J. Med. 372, 2097–2107.

Yu, J., Chen, Z., Zhang, T., Li, Zhiling, Ni, Y., Li, Zhongxiang, 2011. Association of genetic variants in CFTR gene, IVS8 c.1210-12T[5_9] and c.1210-35_1210-12GT[8_12], with spermatogenetic failure: Case-control study and meta-analysis. Mol. Hum. Reprod. 17, 594–603.

Yu, X.C., Li, M.J., Cai, F.F., Yang, S.J., Liu, H. Bin, Zhang, H.B., 2021. A new TEX11 mutation causes azoospermia and testicular meiotic arrest. Asian J. Androl. 23, 510–515.

Zhang, Q., Ji, S.Y., Busayavalasa, K., Yu, C., 2019. SPO16 binds SHOC1 to promote homologous recombination and crossing-over in meiotic prophase I. Sci. Adv. 5, eaau9780.
